# Modelling Country-Level Trajectories of Antimicrobial Resistance in Human Populations

**DOI:** 10.1101/2025.11.20.25340582

**Authors:** Oraya Srimokla, Sean M. Cavany, James Petrie, Mark Pritchard, Anna Nebykova, Gisela R. Aguilar, Paul Turner, Carlo Perrone, Sara Chandy, Tamalee Roberts, Victor D. Rosenthal, Rumina S. Hasan, Nidanuch Tasak, Christiane Dolecek, Ben S. Cooper

## Abstract

Antimicrobial resistance (AMR) is recognised as a global health threat but temporal trends in the prevalence of resistance are typically obscured by patchy and heterogeneous data, particularly in lower and middle-income settings.

Previous approaches to estimating country-level resistance trajectories in such settings have used an ensemble approach, combining multiple flexible nonlinear algorithmic mean functions embedded within a Gaussian process framework without explicitly accounting for heterogeneities between data sources. We build on this work, but aim to overcome important limitations by explicitly accounting for variation between data sources and using a semi-mechanistic approach based on a logistic growth model with time-varying parameters. We take as an example *Acinetobacter baumannii*, a globally significant cause of antibiotic-resistant healthcare-associated infections and use data from 18 different sources to estimate trajectories in the prevalence of resistance in 32 countries in Asia between 2000 and 2022.

Working within a Bayesian framework, we compare six variants of the underlying model which vary in how data source heterogeneities are accounted for and in how the growth model parameters are assumed to vary by country and time and to depend on covariates and spatial relationships. We also consider a stacked ensemble of these six models. We assess out of sample model accuracy using the Leave-One-Out Cross-Validation Information Criterion.

For four out of seven pathogen-antibiotic combinations considered, the stacked model had the highest out of sample prediction accuracy. For the other pathogen-antibiotic combinations, models that accounted for heterogeneity between data sources always performed better than models that did not. This framework improves upon existing approaches by offering a biologically plausible structure that can quantify the effects of covariates on resistance dynamics, effectively handle heterogeneous data sources and can be readily extended to incorporate more complex biologically motivated growth processes. This methodology provides a potentially valuable foundation for better understanding AMR resistance dynamics and evaluating public health interventions.

**Author Summary:** Understanding antimicrobial resistance trajectories and their drivers across countries and over time is important for quantifying the changing health burden of antimicrobial resistance, exploring the potential impact of interventions, and for setting control priorities and informing interventions. However, it is challenging because available surveillance data is sparse and comes from sources that differ in both microbiological practices (e.g. who is sampled and when) and the patient groups and facility types that are represented. We developed a way to model country-level resistance trajectories that accounts for these differences between data sources, represents changes in resistance using a biologically-informed logistic growth model, and that allows resistance trajectories for individual countries to be estimated by borrowing strength from other countries, accounting for spatial dependencies. We applied our method to 18 sources of *Acinetobacter baumannii* resistance data from 32 countries in Asia from 2000 to 2022, considering six different model variants as well as an ensemble approach using Bayesian stacking.

Our results show that accounting for variability between data sources improves how accurately models estimate the proportion of drug-resistant isolates for out-of-sample data. Models that accounted for differences between data sources outperformed other models that did not across all seven pathogen-antibiotic combinations. Combining model variants into a single ensemble model produced the most accurate resistance forecasts for most antibiotic combinations when predicting held-out data from the most recent four years, as measured by the mean absolute error. For countries with sparse or absent resistance data, the spatial models generated plausible resistance estimates that reflected trajectories observed in neighbouring countries. Our approach provides a potentially valuable pathway for better understanding resistance trajectories and assessing the potential impacts of different public health interventions.

## 1 Introduction

Antimicrobial resistance (AMR) threatens global health by compromising infection treatment and straining health systems worldwide [1–4]. AMR occurs when microorganisms such as bacteria, viruses, parasites, and fungi develop resistance against antimicrobial medications, such as antibiotics, making infections more difficult to treat [3]. In response to the growing threat of AMR, international collaborations have been established to coordinate global action. The 2015 Global Action Plan (GAP) on AMR [5] led to the institution of AMR surveillance systems such as the WHO Global Antimicrobial Resistance and Use Surveillance System (GLASS) [6], the publication of national targets, and the creation of national action plans [7]. The 2024 UN General Assembly political declaration on AMR further strengthened these commitments [8]. Despite these efforts, significant challenges remain in understanding AMR trajectories [9].

Mathematical models have become essential tools for understanding AMR dynamics and can be broadly classified as phenomenological or mechanistic. Phenomenological models describe relationships in data without making assumptions about the underlying biology or transmission mechanisms [10]. For example, Browne et al. used a spatiotemporal stacked ensemble approach to describe the relationship between a set of covariates and resistance prevalence for *S*. Typhi and *S*. Paratyphi A in 75 endemic countries from 1990 to 2019 [11]. While strengths of this approach include its flexibility and apparent good performance for disease systems close to equilibrium [12], a limitation is that in settings which may be far from equilibrium resistance prevalence estimates may be produced that lack biological plausibility and are inconsistent with background domain-specific knowledge. Furthermore, because this approach does not account for intrinsic dynamics or represent processes driving non-equilibrium changes in resistance, it does not lend itself to the exploration of counterfactual scenarios or long-term forecasting. In contrast, mechanistic models aim to explicitly describe transmission dynamics through simplified representations of known processes and include model parameters with biologically meaningful interpretations [10]. For example, Kachalov et al. developed a compartmental model to represent the spread of carbapenem-resistant *Klebsiella pneumoniae* and extendedspectrum beta-lactamase-producing (ESBL) *Klebsiella pneumoniae* in community and hospital settings in Europe [13], while similar modelling approaches have been used to evaluate the effect of antimicrobial stewardship interventions [14–17]. Both modelling approaches face substantial challenges when applied to global AMR data. Resistance data typically originate from various data sources, including clinical settings, national surveillance programs, and site-specific studies, each with distinct sampling protocols, reporting practices, and data processing standards [3, 16, 18–20]. Moreover, microbiological sampling practices can vary substantially between settings and are thought to have a significant impact on the directly observed prevalence of resistance [21]. Additional challenges include sparse data coverage, particularly in low and middle-income countries, bias in available data towards healthcare-seeking populations, and low blood culture utilisation rates [16, 21–24].

The approach we propose here can be thought of as a semi-mechanistic model. This represents a hybrid approach, sitting between fully mechanistic models, where the underlying dynamical model is a simplified representation of known processes, and phenomenological models, where nothing about the data generation process is assumed. Our mechanistic component builds on the work of Emons et al. [25], who considered European AMR data for 887 pathogen-drug combinations. While they observed that for many pathogen-drug combinations resistance remained stable over two decades, many other resistance trajectories could be well-described by a scaled logistic growth model with an initial exponential increase in resistance followed by stabilisation at an intermediate level. This scaled logistic growth model represents the solution to a simple mechanistic model for resistance where the dynamics of drug-resistant phenotypes are decoupled from those of drug-sensitive phenotypes. We model the relationship between the parameters of this underlying mechanistic model and covariates phenomenologically, without imposing assumptions about mechanisms or even directions of effect.

Our modelling framework also builds on the approach of Bhatt et al. who used a stacked ensemble approach to combine multiple nonlinear models for the mean function of a Gaussian process regression applied to *Plasmodium falciparum* prevalence data in sub-Saharan Africa [12, 26]. However, while Bhatt et al. used a variety of functions to directly map covariates to observed prevalence, in contrast we map covariates to parameters of a dynamic process at a particular place and time; the system’s intrinsic dynamics then determine predicted resistance prevalence for any given country and time.

In this paper we first describe the example data we are considering. We then show how the logistic model emerges as a solution to a simple mechanistic model of resistance transmission, and describe how this basic model structure can be extended across six model variants to incorporate country and source-level variation, spatial correlation, and time-varying covariates that potentially impact resistance dynamics. Following this, we describe model fitting within a Bayesian framework, model performance evaluation using leave-one-out cross-validation, and the application of Bayesian model stacking to generate an ensemble model optimised for out-of-sample predictions. Finally, we present results comparing model performance across seven *Acinetobacter baumannii*-antibiotic combinations in Asia from 2000 to 2022, and discuss the implications of our findings for AMR surveillance and the potential for future public health intervention evaluation.

## 2 Methods

### 2.1 Data

Resistance data for *Acinetobacter baumannii* was obtained from the Global Research on Antimicrobial Resistance 2024 global burden study [4] which makes use of around 1.3 million clinical isolates for 113 countries globally from 1991 to 2022, combining data from surveillance programs, individual testing facilities, and systematic reviews. We extracted resistance data for seven classes of antibiotics: aminoglycosides, antipseu-domonal penicillins, beta-lactams/beta-lactamase inhibitor combinations, carbapenems, fluoroquinolones, fourth-generation cephalosporins, and third-generation cephalosporins. For this analysis, we focus on countries in Asia and the period from 2000 to 2022, for which we retrieved data from 333936 clinical isolates across 18 data sources, 32 countries, and 7 pathogen-antibiotic combinations. See Section A in S1 Text for the total isolates per year from 2000 to 2022 across Asia and the data sources used in this analysis.

Covariate data were selected from previous studies identifying possible associations with resistance in *Acinetobacter baumannii* [27–31]. Covariate data included in the models are time-dependent and country-level gridded mean temperature data from the Copernicus Climate Change Service [32], estimates of total human antibiotic consumption (defined daily doses per 1000 inhabitants per day (DDD per 1000)) and estimates of the fraction of out-of-pocket healthcare expenditures from the 2024 Global Burden of Resistance study [4, 33]. All covariates included were available at year and country level.

### 2.2 Model Structure

Our underlying model of resistance dynamics is a logistic growth process and we consider variants of this basic model with parameters that vary in space and time as a function of covariates. There are a number of possible parameterisations of this model. In this paper, we focus on a parameterisation where the logistic curve emerges as the solution to a two-compartment ordinary differential equation, the susceptible-infectious-susceptible (SIS) transmission model. This SIS model can be expressed as

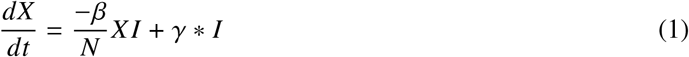

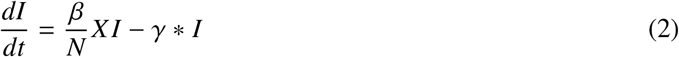

where *X* represents the number susceptible to infection and *I* the number currently infectious.

Mechanistically, we might think of this model as a simple approach for representing carriage dynamics for variants of a particular bacterial species that are resistant to a particular antibiotic when the dynamics of drug-resistant and drug-sensitive strains are decoupled from each other (see Section B in S1 Text ), an assumption that has some empirical support for a number of important nosocomial bacterial pathogens [34]. Under this assumption, the proportion of the population colonised with resistant strains, *P*_*t*_, follows logistic growth dynamics [25], described by the Verhulst equation:

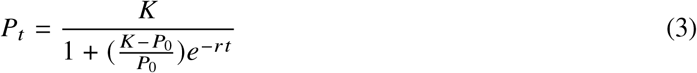

where *K* is the carrying capacity, *r* is the growth rate, and *P*_0_ is the initial proportion colonised. We parameterise this equation in terms of *β* and *γ*, rather than the standard carrying capacity and growth rate. In this parameterisation, the initial exponential growth rate, *r* can be defined as *β* − *γ* and the carrying capacity as (1 − *γ*/*β*) or, equivalently, 1 − 1/*R*_0_, where *R*_0_ is the basic reproduction number (see Section B in S1 Text). Note that, *P*_*t*_ cannot be directly equated to the observed proportion of clinical infections of a particular pathogen that are resistant. This latter quantity depends on additional factors including the proportion of people colonised with drug-sensitive strains, the risk of infection given colonisation (which may differ for resistant and sensitive strains), and the relative chances of drug-resistant and drug-sensitive infections being detected and reported, all which may vary with data. To account for these factors we introduce a source-specific scaling factor, η_*s*_, described below.

We extend this simple model to allow parameters governing the growth process to vary between countries, source, and over time to capture temporal changes in resistance dynamics. For models incorporating covariates, parameters can vary by country and year (see Table 1). For models without covariates, each country has country-specific but temporally constant values for *β* and *γ* and produces a singular growth curve per country. Six model variants are explored with different combinations of country and data source-level variation, spatial correlation effects, and covariates (see Table 2).

**Table 1.** Definitions of model parameters for six model variants. Parameters are defined based on logistic growth dynamics where *P*_*c,t*_ represents the proportion of resistant isolates at time *t* and country *c*. Here Model C: Model with country-level random effects, C+X: Country-level random effects model and covariates, C+S: Country-level random effects and source-level scaling, SP: Spatial model with Gaussian process regression, SP+S: Spatial model with source-level scaling, SP+S+X: Spatial model with source-level scaling and covariates.

| Model | Rate of Resistance Coefficient ($\beta$) | Loss of Resistance Rate ($\gamma$) | Initial Prevalence ($P_0$) | Source Effect ($\eta_s$) |
| --- | --- | --- | --- | --- |
| **Model C** | $\beta_c$<br>(Country-specific) | $\gamma_c$<br>(Country-specific) | $P_{0,c}$<br>(Country-specific) | None<br>($\eta_s = 1$) |
| **Model C + X** | $\beta_{c,t} = \exp(\log \beta_c + \sum \theta_k \cdot X_{k,c,t})$<br>(country-specific+time-varying covariates) | $\gamma_c$ | $P_{0,c}$ | None<br>($\eta_s = 1$) |
| **Model C + S** | $\beta_c$ | $\gamma_c$ | $P_{0,c}$ | $P_{c,s,t}$<br>$= P_{c,t} \times \eta_s$ |
| **Model SP** | $\beta_c = \exp(\log \beta + k_{\beta,c})$<br>(spatial effect $k_{\beta}$) | $\gamma_c = \exp(\log \gamma + k_{\gamma,c})$<br>(spatial effect $k_{\gamma}$) | $P_{0,c} = \text{logit}^{-1}(P_{0,\text{logit}} + k_{P_{0,c}})$<br>(spatial effect $k_{P_0}$) | None<br>($\eta_s = 1$) |
| **Model SP + S** | $\beta_c = \exp(\log \beta + k_{\beta,c})$ | $\gamma_c = \exp(\log \gamma + k_{\gamma,c})$ | $P_{0,c} = \text{logit}^{-1}(P_{0,\text{logit}} + k_{P_{0,c}})$ | $P_{c,s,t}$<br>$= P_{c,t} \times \eta_s$ |
| **Model SP + S + X** | $\beta_{c,t} = \exp(\log \beta + k_{\beta,c} + \sum \theta_k \cdot X_{k,c,t})$<br>(spatial effect $k_{\beta}$ + time-varying covariates) | $\gamma_c = \exp(\log \gamma + k_{\gamma,c})$ | $P_{0,c} = \text{logit}^{-1}(P_{0,\text{logit}} + k_{P_{0,c}})$ | $P_{c,s,t}$<br>$= P_{c,t} \times \eta_s$ |

**Table 2.** Model variants and the model components that make up each model. Here Model C: Model with country-level random effects, C+X: Country-level random effects model and covariates, C+S: Country-level random effects and source-level scaling, SP: Spatial model with Gaussian process regression, SP+S: Spatial model with source-level scaling, SP+S+X: Spatial model with source-level scaling and covariates.

|  | Model C | Model C + X | Model C + S | Model SP | Model SP + S | Model SP + S + X |
| --- | --- | --- | --- | --- | --- | --- |
| Country-level Random Effects | ✓ | ✓ | ✓ |  |  |  |
| Covariates |  | ✓ |  |  |  | ✓ |
| Data source-level Random Effects |  |  | ✓ |  | ✓ | ✓ |
| Spatial Random Effects |  |  |  | ✓ | ✓ | ✓ |

The proportion of the population colonised with resistant strains for the most general model (SP+S+X) at year *t* for country *c* and source *s* can be defined by the following piecewise logistic function:

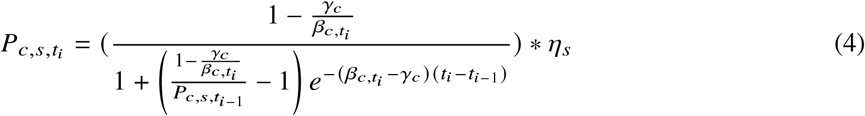

where 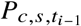 is the country- and source-specific resistance prevalence from the previous observation at index *i* − 1, η_*s*_ is a source-specific multiplicative scaling factor, and consecutive times *t*_*i*−1_ and *t*_*i*_ can be unequally spaced. After model fitting, country-specific trajectories are generated for each year from 2000 to 2022 from fitted model objects, later described below. The initial condition, 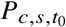 is defined as:

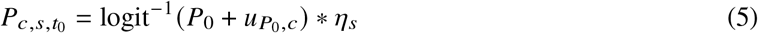

where *P*_0_ is the population-level initial resistance value in the logit space, and 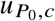 is the spatial random effect for the initial prevalence of resistance for country *c*.

For this model, we represent 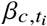 as a function of time- and country-specific covariates:

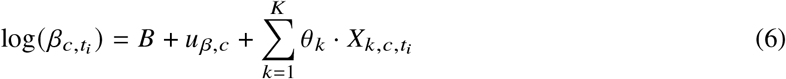

where *B* is an intercept term, 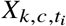 are the K normalised covariates for country *c* at year *t*_*i*_, *u*_*β,c*_ represents the spatial effect for *β*_*c*_ for country *c*, and *θ*_*k*_ are the estimated covariate coefficients (see Section C and Section D in S1 Text ). This structure ensures that 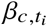 remains positive while allowing time-varying dynamics through the addition of covariates. For simplicity, we assume here that *γ*_*c*_ remains constant:

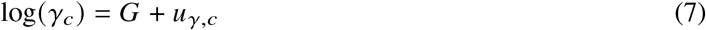

where *G* is an intercept term, and *u*_*γ,c*_ is the spatial effect for *γ*_*c*_ for country *c* ( see Table 1).

Spatial effects are incorporated through Gaussian process regression to capture spatial autocorrelation between neighbouring countries [35, 36]. To establish relationships between geographically close sites, we use a radial basis kernel function to define the covariance between countries according to the scaled distance (see Section D in S1 Text ):

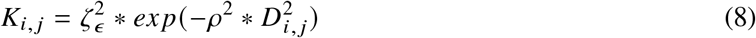

where *D*_*i, j*_ is the scaled Euclidean distance from geographic centroids of countries *i* and *j*, 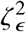 is the maximum covariance between sites for model parameter *ϵ*, and *ρ*^2^ is the rate at which covariance declines with distance. This covariance function is used to estimate spatially-correlated random effects for each country and is used to define 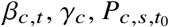 [35, 36].

Simpler nested model variants are obtained by constraining components of the general model (SP+S+X). For model variants that retain spatial random effects, removing covariates gives a spatially-varying but temporally constant *β*_*c*_ and source-level scaling (SP+S). Further removing source-level scaling (η_*s*_ = 1) gives a model with only spatial random effects (SP).

Country-level random effects allow *β* and *γ* to vary by country *c* with hierarchical priors (see Table 1 and Section E in S1 Text ). The spatial structure can alternatively be replaced by country-level random effects by removing spatial effect terms *u*_*β,c*_ and *u*_*γ,c*_, with intercept terms *B* and *G* instead drawn from a shared population-level distribution:

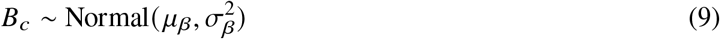

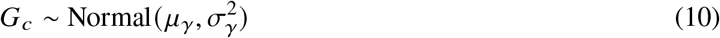

where *μ*_*β*_ and *μ*_*γ*_ are the population-level means and 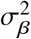 and 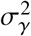 are the between-country variances (see Section E in S1 Text). Three model variants are obtained with country-level random effects: country-level effects with only source-level scaling (C+S), country-level effects with only covariates (C+X), and the simplest variant, country-level effects with no covariates and source-level scaling (C) (see Table 1 and Section E in S1 Text) .

Observations of resistant isolates are assumed to follow a binomial distribution:

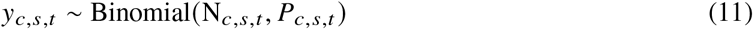

where *N*_*c,s,t*_ is the total number of isolates tested for a given country-source combination at time *t*, and *P*_*c,s,t*_ is the predicted proportion of clinical isolates from sources which are resistant.

#### 2.2.1 Countries with Missing Data

For countries without resistance data, the Gaussian process framework enables spatial imputation using the posterior estimates of the maximum covariance between sites *ζ*^2^, and the rate at which covariance declines with distance *ρ*^2^. New values are predicted following a multivariate normal predictive distribution:

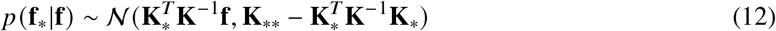

where *f* is the observed spatial effects for countries with resistance data, *f*_*_ is the spatial effects for countries with no resistance data, *K* is the covariance matrix of countries with resistance data, *K*_*_ is the covariance matrix with a combination of countries with and without resistance data, and *K*_**_ is the covariance matrix of countries with no resistance data [37, 38].

#### 2.2.2 Ensemble Modelling

To combine predictions across model variants, we apply Bayesian model stacking using the loo (v2.8.0) package in R [39]. Model stacking generates ensemble predictions by finding the combination of models that maximises the leave-one-out (LOO) predictive density and the expected log pointwise predictive density (ELPD) of the ensemble predictions [12, 26, 40]. LOO predictive densities are obtained using Pareto-smoothed importance sampling leave-one-out cross-validation (LOO-CV), a method for estimating pointwise out-of-sample prediction accuracy from a fitted Bayesian model [39]. The optimal model weights for each pathogen-antibiotic combination were obtained using the 100 model weights() function, and used to determine the contribution of each model’s predictive distribution to the ensemble [26, 39].

### 2.3 Model Fitting and Evaluation

Models were implemented in the Stan programming language, and model fitting and Hamiltonian Monte Carlo (HMC) sampling performed via the RStan interface (version 2.32.6).

Prior distributions for the models were selected to be weakly informative or informative when domain knowledge suggested biologically plausible parameter ranges (see Section C to F in S1 Text ). We conducted prior and posterior predictive checks. For prior predictive checks, data were simulated from prior distributions to assess if the priors produce reasonable model outputs [41]. After fitting the model to empirical data, posterior predictive checks were done by simulating new replicated data sets based on the fitted model [41]. For each of the models, four chains were run with 5000 iterations per chain, of which 2500 were warmup. We assessed convergence using standard MCMC diagnostics including traceplots and Rhat statistics (with Rhat < 1.01). We additionally examined the number of divergent transitions, maximum tree depth, and pairs plots to identify issues with convergence or model parameterisation. Parameter recoverability was assessed by fitting the models to simulated data with known parameter values.

The models were then fitted with empirical data. For each of the six model variants, posterior parameter estimates, observation-level predictions, and country-specific growth curves for the years 2000 to 2022 were extracted from fitted model objects.

The performance of the six individual model variants (see Table 2) for each pathogen-antibiotic combination were compared using the Leave-One-Out Cross-Validation Information Criterion (LOOIC) obtained from applying the LOO-CV method via the loo (v2.8.0) package in R [42]. The LOOIC value was computed from the ELPD, with lower LOOIC values indicating better expected out-of-sample predictive performance. To evaluate the models’ performance for forecasting future resistance trends we fitted models to a training dataset and evaluated the predictions on a test set created by leaving out the most recent four years of data for each country. Only countries with at least one observation in the final four years contributed to the test set. Predictive accuracy for this assessment was quantified using the mean absolute error (MAE) between observed and predicted values.

## 3 Results

After fitting model variants to all countries in Asia from 2000 to 2022 we found that all six model variants estimated qualitatively similar temporal trajectories for resistance for each country, though for countries with sparse data in the early 2000s, spatial models produced trajectories more influenced by neighbouring countries (see Fig 1 and Section G in S1 Text). In most cases (192 of 214 antibiotic-country combinations) increasing resistance trends were estimated, though in a few cases (22 of 214 antibiotic-country combinations) declining trends were seen, most notably in Japan for all seven antibiotic combinations. Using models without covariates (C, C+S, SP, SP+S), we estimated the long-term saturation levels of resistance for each country and found that the majority of countries showed intermediate saturation levels rather than resistance fixation (see Section H in S1 Text). For models with covariates, we observe year-to-year fluctuations in temporal resistance trajectories as a result of time-varying covariates (see Section G in S1 Text). As expected, uncertainty in estimated resistance proportions varied substantially by country and time, with high levels of uncertainty typically seen at the beginning of the time series where the data were most sparse. There was also substantial variation in levels of uncertainty in the estimated prevalence of resistance between the different models, with the spatial models (SP, SP+S, SP+S+X) typically giving narrower predictive intervals for country-level resistance proportions when data was sparse, a consequence of borrowing information from nearby countries.

**Fig. 1.**
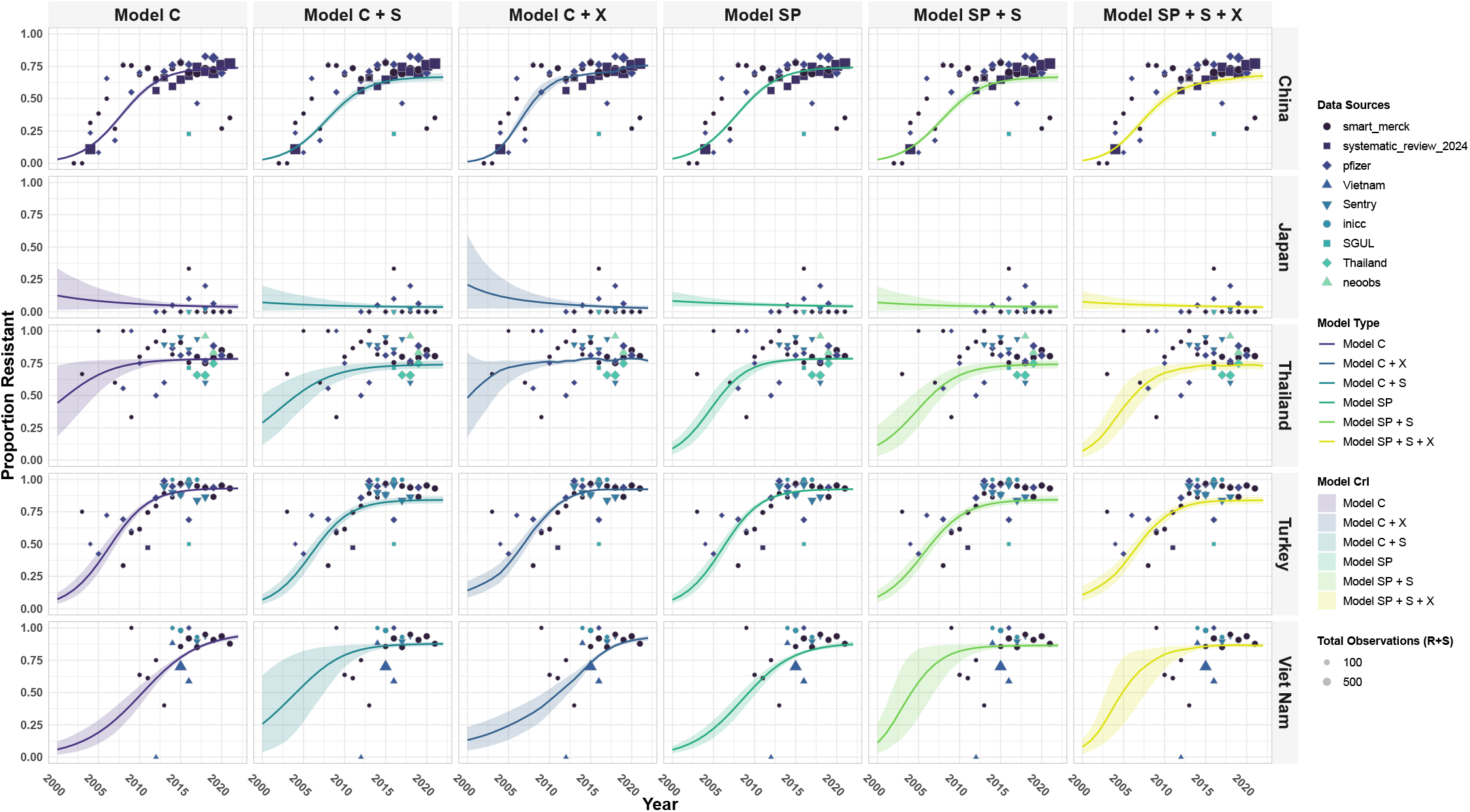
Individual model fits using *β, γ, P*_0_ parameterisation of the logistic growth function for carbapenem-resistance in *Acinetobacter baumannii* for five countries in Asia. Posterior predictions for the proportion of clinical isolates that are resistant to carbapenems for five countries and six model variants. See Table 2 for model definitions. Solid lines represent the mean posterior prediction and the shaded regions show the 95% credible intervals with the different colours corresponding to each model type. The points show the observed resistance data used for training the models.

Comparing out-of-sample predictive performance using leave-one-out cross-validation, in all cases, models that included random effects showed substantially lower LOOIC values (indicating better performance) than models that did not ( Table 3). Comparing models that included covariates with corresponding models that did not (C+X v C and SP+S v SP+S+X) we found that including covariates consistently improved predictive performance for five out of the seven antibiotic categories considered (aminoglycosides, antipseudomonal penicillins, carbapenems, fluoroquinolones and third-generation cephalosporins), and in some cases led to substantial improvements. The model incorporating both country-level and source-level scaling (C+S) produced the lowest LOOIC value across five of seven antibiotic categories, indicating the best predictive performance (see Table 3). For the remaining two antibiotic categories (aminoglycosides and antipseudomonal penicillins), the model combining spatial effects and source-level scaling with covariates (SP+S+X) had the lowest LOOIC. Model C with only country-level random effects consistently had the second highest LOOIC, while the base spatial model (SP) had the highest LOOIC values for six of seven antibiotic categories. These results suggest that country-level effects and spatial effects alone are not able to capture heterogeneity in the resistance data. In contrast, spatial models incorporating source-level scaling and/or covariates had relatively good predictive performance, ranking in the top three across all antibiotic combinations.

**Table 3.** Comparison of fitted models for each antibiotic class using the leave-one-out cross-validation information criterion (LOOIC). Relative differences in LOOIC calculated from the best model (shown in bold with 0). Lower values indicate better predictive performance. See Table 2 for model definitions.

| Antibiotic<br>Class/Subclass | Model Comparison using LOOIC differences from best model |  |  |  |  |  |
| --- | --- | --- | --- | --- | --- | --- |
|  | Model<br>C | Model<br>C+X | Model<br>C+S | Model<br>SP | Model<br>SP+S | Model<br>SP+S+X |
| **Aminoglycosides** | 431.3 | 430.4 | 38.3 | 466.9 | 4.9 | **0** |
| **Antipseudomonal**<br>**penicillins** | 200.8 | 136.6 | 49.2 | 221.0 | 71.8 | **0** |
| **Beta-lactams/beta-**<br>**lactamase inhibitor**<br>**combinations** | 196.0 | 210.0 | **0** | 230.0 | 55.4 | 47.3 |
| **Carbapenems** | 378.2 | 329.9 | **0** | 432.2 | 100.9 | 95.8 |
| **Fluoroquinolones** | 663.6 | 567.7 | **0** | 722.9 | 27.7 | 13.3 |
| **Fourth-generation**<br>**cephalosporins** | 131.9 | 132.7 | **0** | 130.8 | 31.7 | 30.7 |
| **Third-generation**<br>**cephalosporins** | 312.2 | 221.9 | **0** | 334.3 | 56.6 | 9.5 |

An ensemble modelling approach using Bayesian model stacking has potential to improve predictive performance. The model stacking weights are shown in Table 4. The model incorporating both country-level and source-level scaling (C+S) was assigned the highest weight for five out of seven antibiotic categories, indicating optimal prediction accuracy if the sampling is heavily weighted for this model. For the remaining two antibiotics categories, aminoglycosides and antipseudomonal penicillins, the highest weights were assigned to the spatial model with source-level scaling (SP+S) and the country-level effects model with covariates, respectively. The model with only country-level random effects (C), and/or the model with only spatial effects (SP) generally received lower weightings across different antibiotic categories. Examples of stacked model predictions for nine countries using these stacking weights are shown in Fig 2.

**Table 4.** Comparison of fitted models weights for each antibiotic category using model stacking. The models with the highest weights are shown in bold. See Table 2 for model definitions.

| Antibiotic<br>Class/Subclass | Model Stacking Weights |  |  |  |  |  |
| --- | --- | --- | --- | --- | --- | --- |
|  | Model<br>C | Model<br>C+X | Model<br>C+S | Model<br>SP | Model<br>SP+S | Model<br>SP+S+X |
| Aminoglycosides | 0.16 | 0.12 | 0.04 | 0.10 | **0.32** | 0.26 |
| Antipseudomonal<br>penicillins | 0.03 | **0.34** | 0.12 | 0.02 | 0.26 | 0.23 |
| Beta-lactams/beta-lactamase inhibitor<br>combinations | 0.08 | < 0.001 | **0.63** | 0.13 | 0.06 | 0.10 |
| Carbapenems | < 0.001 | 0.29 | **0.48** | 0.16 | 0.05 | 0.02 |
| Fluoroquinolones | 0.02 | 0.18 | **0.44** | 0.02 | 0.28 | 0.06 |
| Fourth-generation<br>cephalosporins | < 0.001 | 0.06 | **0.54** | 0.23 | < 0.001 | 0.17 |
| Third-generation<br>cephalosporins | 0.02 | 0.26 | **0.37** | 0.02 | < 0.001 | 0.33 |

**Fig. 2.**
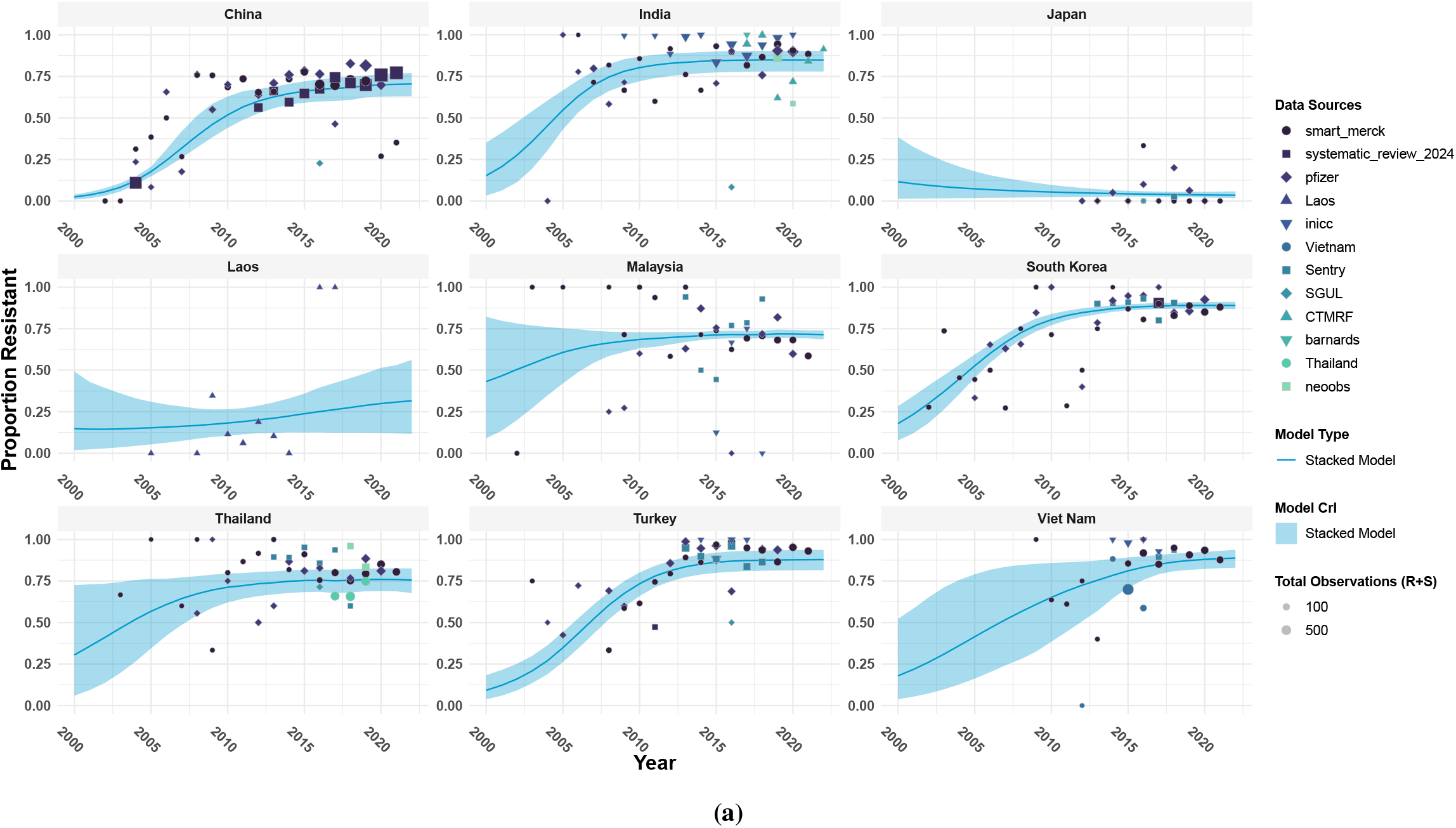
Stacked model predictions using *β γ, P*_0_ parameterisation of the logistic growth function for resistance of *Acinetobacter baumannii* and carbapenems for Asia from 2000 to 2022. Stacked model estimates for nine countries are shown. Solid lines represent the mean posterior prediction and the shaded regions show the 95% credible intervals. The points show the observed resistance data used for training the models.

For models incorporating source-level random effects, the MAE was calculated for each country-source-time combination and for all other models, it was calculated for each country-time combination. The MAE values for all pathogen-antibiotic combinations are shown in Table 5. The stacked model had the lowest MAE value for four out of seven antibiotic categories, while the spatial model with source-level random effects and covariates (SP+S+X) had the lowest for two and the model with country-level random effects and source-level scaling (C+S) had the lowest MAE for a single antibiotic category. Similar to the LOOIC model comparison, models only incorporating country-level random effects (C) or only spatial effects (SP) tended to have lower predictive accuracy. The spatial model (SP) was found to have the largest MAE values for four of seven antibiotic categories.

**Table 5.** Comparison of mean absolute error (MAE) calculated for four years of held-out data across countries, years, and sources (for source-specific models). The MAE was calculated for a held-out test set containing the most recent four years of data for each country. The best model for each antibiotic category is shown in bold. Lower MAE values indicate more accurate model predictions. See Table 2 for model definitions.

| Antibiotic<br>Class/Subclass | Mean Absolute Error |  |  |  |  |  |  |
| --- | --- | --- | --- | --- | --- | --- | --- |
|  | Model<br>C | Model<br>C+X | Model<br>C+S | Model<br>SP | Model<br>SP+S | Model<br>SP+S+X | Stacked<br>Model |
| Aminoglycosides | 0.1684 | 0.1672 | 0.1525 | 0.1646 | 0.1513 | **0.1511** | 0.1532 |
| Antipseudomonal<br>penicillins | 0.1284 | 0.1277 | 0.1222 | 0.1294 | 0.1257 | 0.1236 | **0.1210** |
| Beta-lactams/beta-lactamase inhibitor<br>combinations | 0.1861 | 0.1854 | 0.1872 | 0.1848 | 0.1890 | **0.1847** | 0.1951 |
| Carbapenems | 0.1406 | 0.1403 | 0.1362 | 0.1423 | 0.1392 | 0.1374 | **0.1361** |
| Fluoroquinolones | 0.1187 | 0.1188 | 0.1215 | 0.1250 | 0.1215 | 0.1203 | **0.1177** |
| Fourth-generation<br>cephalosporins | 0.1288 | 0.1302 | 0.1231 | 0.1273 | 0.1219 | 0.1221 | **0.1216** |
| Third-generation<br>cephalosporins | 0.1333 | 0.1340 | **0.1259** | 0.1429 | 0.1367 | 0.1370 | 0.1267 |

Analysis using simulated data with the country-level random effects model with a *β*_*c*_, *γ*_*c*_, and *P*_0,*c*_ parameterisation confirmed that the models were able to recover the true values of the parameters; posterior predictive checks further confirmed the model was able to capture the mean and standard deviation of the observed data (see Section J in S1 Text ). The convergence of all six model variants was visually checked for all seven pathogen-antibiotic combinations using trace plots and pairs plots (see Section K in S1 Text ). All plots exhibit convergence of all four chains in the trace plot, appropriate Rhat values, and do not show model divergences.

## 4 Discussion

We developed a Bayesian modelling framework that is able to account for time-varying parameters, country and source-level variation, covariates, and spatial correlation effects to estimate country-level antimicrobial resistance trajectories. We compared six model variants applied to seven *Acinetobacter baumannii*-antibiotic combinations across Asia from 2000 to 2022 and explored the applications of model stacking to generate ensemble predictions. We found that models incorporating source-level scaling consistently outperformed other models, as shown by the lowest LOOIC values. We accounted for spatial correlation effects in some of our models and observed an improvement in model performance when including covariates in the spatial and source-level scaling model (SP+S). We found that in most cases the ensemble approach, combining estimates from multiple component models, led to improved predictive accuracy. This framework is able to capture resistance dynamics of AMR across different countries, and enables estimates for countries where data are sparse by borrowing information from other countries.

While our modelling approach is broadly phenomenological in the sense that we aim to capture the sigmoidal growth process that changes in AMR prevalence typically follow without making explicit mechanistic assumptions, we note that the models with source-level scaling can be given a semi-mechanistic interpretation as the solution of a deterministic SIS model for the resistant strain, assuming no interaction of drug-resistant and drug-sensitive strain dynamics and a constant prevalence of drug-sensitive strains. This simple model might be considered an appropriate mechanistic representation of the transmission process when there is minimal niche overlap between drug-resistant and drug-sensitive strains [43].

The presence of an analytical solution to this system leads to computational efficiency making it an appealing choice compared to alternative models of sigmoidal growth when used as the basis for complex models applied to multicountry data. The underlying modelling framework we propose, however, can readily be extended to systems corresponding to different mechanistic assumptions. We derived our model in terms of *β* and *γ* as this parameterisation showed improved model convergence when compared alternative parameterisations. Within this framework we included time-varying parameters based on non-linear mappings between covariates that were thought likely to influence resistance dynamics such as antibiotic usage and hospital resourcing levels while accounting for spatial correlation and heterogeneity between data sources.

The framework presented is well-suited to capturing resistance dynamics where, as observed by Emons et al. across European pathogen-drug combinations, trajectories can reflect a mix of non-equilibrium and shifting equilibrium frequencies in response to changing antibiotic use. This framework has some advantages over alternative approaches that map covariates directly to observed prevalence data thus failing to account for intrinsic resistance dynamics that result from the communicable nature of the pathogens.

Our model comparison revealed that model structures capable of accounting for source-level heterogeneity (which also corresponds to the subset of models amenable to a mechanistic interpretation) had improved predictive accuracy. We found that models incorporating source-level scaling with the country-level effects model or the spatial model had the best performance across all antibiotic categories, shown by the lowest LOOIC. AMR data originate from various sources which have differences in patient care, patient characteristics, local resistance prevalence (within country variation), and microbiological culture practices (sampling protocols and laboratory methods). Accounting for source-level variability implicitly accounts for these differences. Our final model stacking weights similarly reflect the better performance of these models, as the highest weight is assigned to models with data source-level scaling for six out of seven antibiotic categories.

Incorporating covariates generally improved model performance across both models with only country-level effects (C) and the spatial effects model (SP). For the model with only country-level effects (C), adding covariates (C+X) reduced the LOOIC values for five out of seven antibiotic categories. We observed a similar pattern when adding covariates to our spatial models. When comparing the spatial model (SP), the spatial model with data source-level scaling (SP+S), and the spatial model with data source-level scaling and covariates (SP+S+X), we see a decrease in LOOIC values across all antibiotic categories when covariates are added to the spatial model with data-source level effects.

When comparing the base spatial model (SP) to the country-level effects model (C), the addition of spatial correlation effects did not consistently reduce LOOIC values across all antibiotic categories. Similarly, combining spatial effects with source-level scaling (SP+S) did not always improve predictive performance relative to the country effects and data source-level scaling (C+S) model. Nevertheless, incorporating a spatial correlation structure provides a useful feature that enables the estimation of resistance trends for countries without observed resistance data. Through the Gaussian process structure of the spatial model, we can estimate trends for data-sparse countries by borrowing information from their neighbouring countries. Our current implementation uses geographic distance to define the spatial correlation structure and assumes neighbouring countries share similar resistance patterns. However, alternative spatial definitions could be explored within this framework. For example, human movement patterns through travel and migration can be more mechanistically relevant when considering a spatial structure in terms of resistance dynamics [17].

This study has several limitations. Our model parameterisation is derived from a two-compartment system that simplifies the dynamics of drug-resistant and drug-sensitive strains and is not intended to be given a literal mechanistic interpretation. Our example pathogen, *Acinetobacter baumannii*, is primarily a healthcare-associated pathogen. When the current version of the model is applied to pathogens that circulate in both hospital and community settings (e.g. *Staphylococcus aureus*), model predictions may be less accurate as it does not distinguish between these settings or stratify by infection type, which may differ in transmission dynamics, antibiotic selection pressures, and sampling practices. Due to limited availability of hospital-level antibiotic consumption data, the antibiotic consumption covariate used here represents the total human antibiotic consumption at the country level, which may not accurately reflect hospital-level antibiotic pressure relevant for healthcare-associated pathogens such as *Acinetobacter baumannii*. More complex formulations that include hospital and community compartments and separate antibiotic use by hospital and community use and by antibiotic class might be expected to lead to more meaningful mechanistic insights as well as having the potential for better predictive performance. Also, while our framework currently allows for time-varying parameters driven by covariates, both evolutionary and ecological processes and unmeasured covariates could lead to changes in the dynamical system that the current framework does not capture. To address these there may be additional benefits in allowing model parameters to change over time for reasons other than covariate change, for example through the incorporation of a random walk process into the model. Finally, further extensions would be needed to account for potential biases in resistance data resulting from microbiological culture practices [21] and changing resistance breakpoints.

In this work, we developed a Bayesian semi-mechanistic modelling framework designed to address the challenge of estimating AMR dynamics across large spatial scales where data sources are often scarce and heterogeneous. This framework accounts for data source heterogeneity, covariate data, intrinsic resistance dynamics and spatial correlation structures. We found that the best-fitting models were those which both accounted for data source heterogeneity and which lent themselves to a biological interpretation. Using a model stacking approach led to improved predictive performance for most antibiotic combinations. This methodology provides a framework for using diverse data sources to understand the dynamics of global resistance trends and offers the potential for extension to more explicitly mechanistic formulation which would provide a foundation for evaluating potential public health interventions.

## Data Availability

All data produced in the present study are available upon reasonable request to the authors and data collaborators

## Acknowledgements

We would like to thank the GRAM team at the Institute for Health Metrics and Evaluation and Oxford and all our GRAM data collaborators for building the foundation of this project and making this work possible. We would also like to thank the UK Department of Health and Social Care’s Fleming Fund, and the Wellcome Trust for funding this project. We would like to acknowledge the NeoOBS Study Collaborators, Global Antibiotic Research and Development Partnership, Geneva, Switzerland. We acknowledge Cardiff University in relation to use of data from the BARNARDS Study.

## Data Availability

The GRAM Project is committed to enabling appropriate access to its data for external researchers. Data collected in Oxford may be made available through the Antimicrobial Resistance (AMR) data repository hosted by the Infectious Diseases Data Observatory (IDDO) at https://www.iddo.org/antimicrobial-resistance, where contributing collaborators have agreed to deposit their data. Access is provided on request and is subject to review and approval by the Data Access Committee (DAC) or the contributing collaborator, in line with applicable governance frameworks. Data collected by the Institute for Health Metrics and Evaluation (IHME) can be accessed through the Global Health Data Exchange (GHDx) at https://ghdx.healthdata.org/. Publicly available datasets are additionally accessible through the Vivli AMR surveillance data sharing platform at https://amr.vivli.org/. For data not publicly available through these repositories, access is subject to the terms and conditions of the original data contributors. Researchers wishing to request access to these data should contact the corresponding author, who will facilitate introductions to the relevant data contributors. Access will then be considered by the data contributors in accordance with applicable data sharing agreements.

All code used in this study is publicly available on GitHub at https://github.com/orayasrimokla/methods_modelling_trajectories_AMR_manucript

## Supporting Information

### S1 Text. Supporting Model Outputs

Combined file of supporting text and figures.

## Supporting Information

### A Data Summary

Figures to summarise data and the total number of isolates for *Acinetobacter baumannii* by source and by antibiotic class and subclass for Asia from 2000 to 2022. The data used from the GRAM 2024 study compiled intrinsic resistance exclusions from both the CLSI and EUCAST guidelines [4]. For third-generation cephalosporins, cefotaxime and ceftriaxone resistance data for *Acinetobacter baumannii* were excluded based on EUCAST guidelines.

**Fig. 1.**
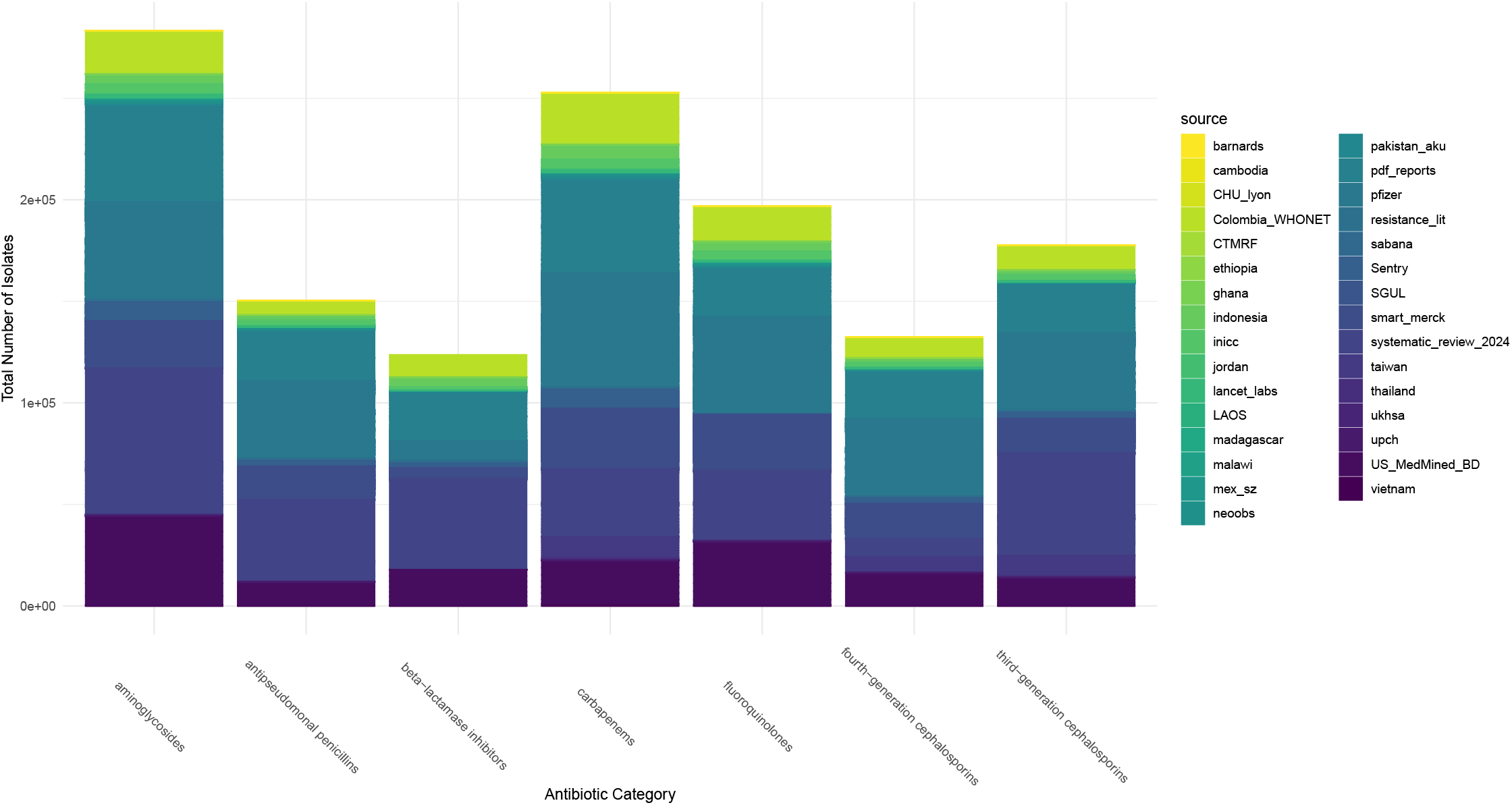
Total number of Acinetobacter baumannii isolates from global datasets. The total number of isolates for 113 countries from 1991 to 2022 are shown across 7 antibiotic categories.

**Fig. 2.**
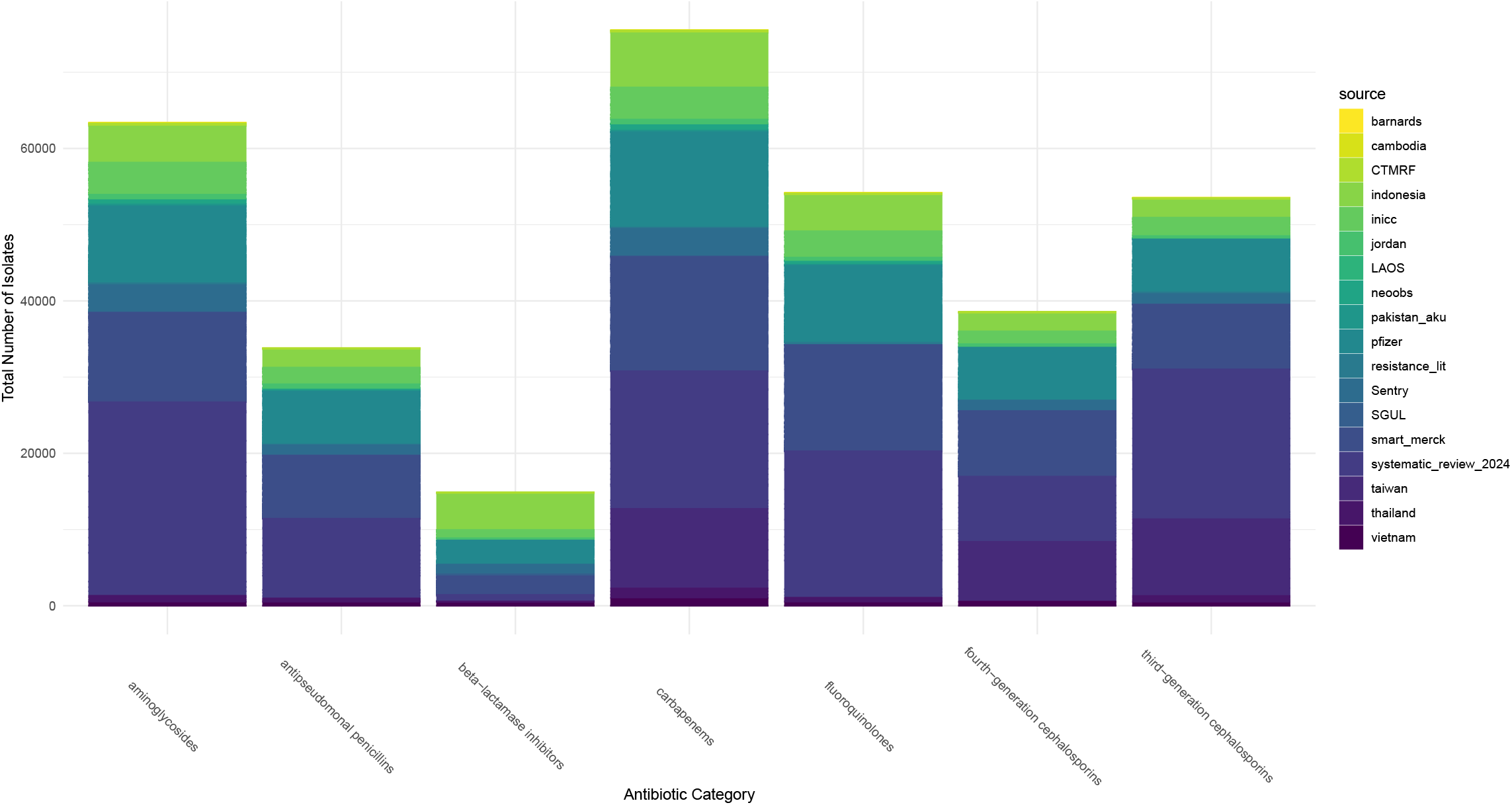
Total number of isolates for Acinetobacter baumannii data across Asia. The total number of isolates for 32 countries in Asia from 2000 to 2022 are shown across 7 antibiotic categories.

**Fig. 3.**
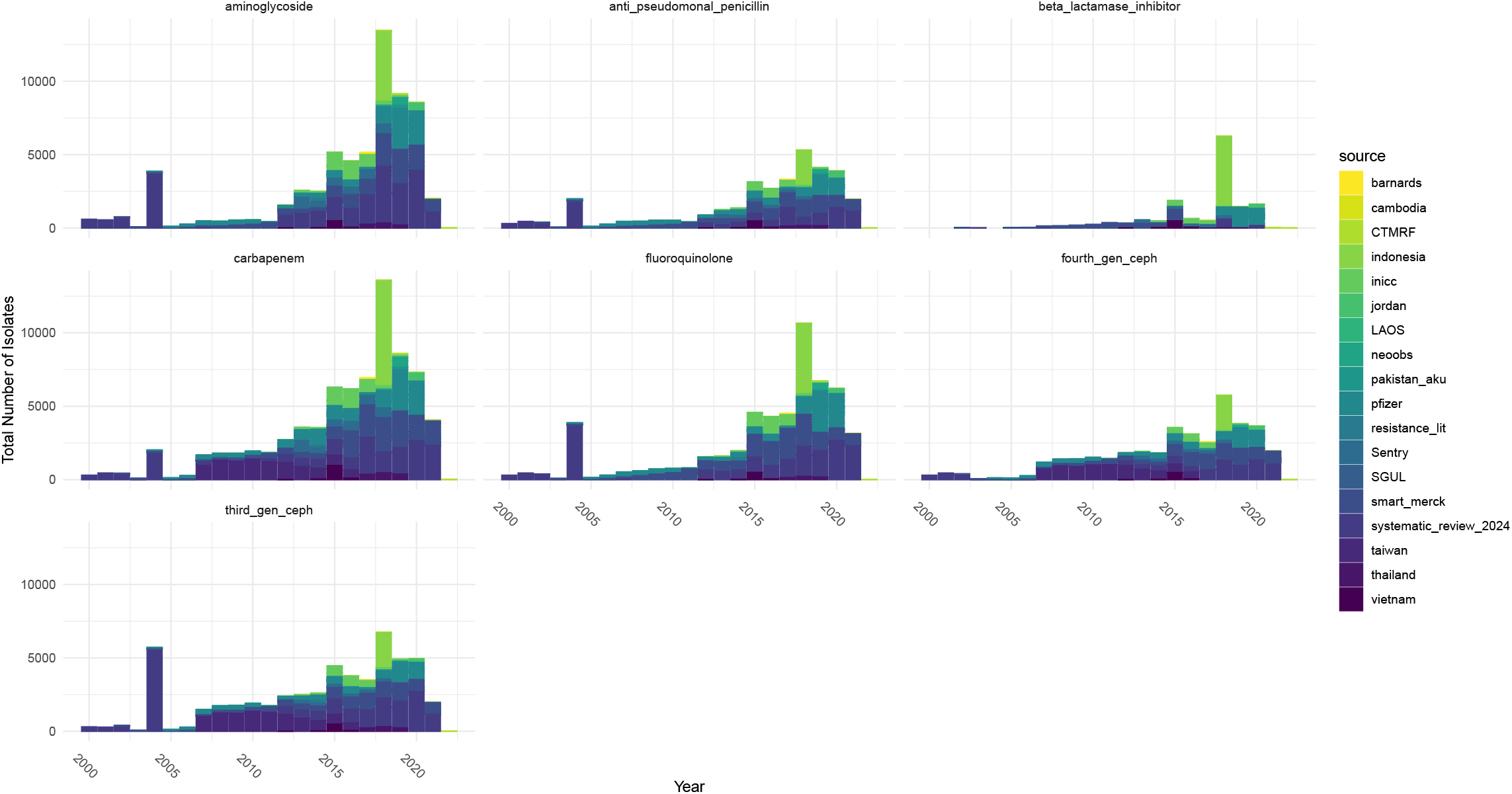
Total number of isolates for *Acinetobacter baumannii* data across Asia per year. The total number of isolates for 32 countries in Asia from 2000 to 2022 are shown across 7 antibiotic categories.

**Table 1.** Total number of isolates for *Acinetobacter baumannii* by source and by antibiotic class/subclass used for Asia from 2000 to 2022.

| Source | Aminoglycosides | Antipseudomonal penicillins | Beta-lactams/beta-lactamase inhibitor combinations | Carbapenems | Fluoroquinolones | Fourth-generation cephalosporins | Third-generation cephalosporins |
| --- | --- | --- | --- | --- | --- | --- | --- |
| CTMRF | 339 | 173 | 177 | 333 | 174 | 167 | 160 |
| LAOS | 106 | – | 2 | 67 | 98 | – | 97 |
| SGUL | 65 | 39 | 13 | 62 | 50 | 22 | 27 |
| Sentry | 3580 | 1344 | 1275 | 3640 | – | 1342 | 1346 |
| BARNARDS | 63 | 21 | – | 42 | 42 | 21 | 21 |
| Cambodia | 63 | – | – | – | 63 | – | 41 |
| Indonesia | 4778 | 2389 | 4768 | 7142 | 4778 | 2389 | 2389 |
| INICC | 4206 | 2177 | 1097 | 4213 | 3449 | 1672 | 2409 |
| Jordan | 530 | 492 | 284 | 543 | 327 | 278 | 240 |
| NeoOBS | 634 | 341 | – | 816 | 502 | 182 | – |
| Pakistan AKU | 245 | 125 | 1 | 222 | 124 | 1 | 123 |
| Pfizer | 10107 | 6941 | 3166 | 12519 | 10107 | 6941 | 6941 |
| Resistance Lit | 235 | 99 | 10 | 160 | 284 | 23 | 227 |
| SMART Merck | 11726 | 8255 | 2678 | 14989 | 13920 | 8590 | 8506 |
| Systematic Review 2024 | 25394 | 10459 | 864 | 18058 | 19206 | 8557 | 19640 |
| Taiwan | – | – | – | 10450 | – | 7854 | 10081 |
| Thailand | 766 | 432 | 21 | 1189 | 512 | – | 749 |
| Vietnam | 550 | 537 | 544 | 1083 | 549 | 537 | 538 |

**Table 2.** Data sources used for *Acinetobacter baumannii* resistance data across Asia from 2000 to 2022.

| Source Label | Full Name | Region/Countries | Years |
| --- | --- | --- | --- |
| Barnards | Burden of Antibiotic Resistance in Neonates from Developing Societies (BARNARDS) | South Asia | 2016–2018 |
| Cambodia | Cambodia Oxford Medical Research Unit and Angkor Hospital for Children Suspected Invasive Bacterial Infection Hospitalizations | Cambodia | 2012–2018 |
| CTMRF | Chennai CHILDS Trust Medical Research Foundation | India | 2018–2022 |
| Indonesia | Indonesian Association of Clinical Pathology and Laboratory Medicine | Indonesia | 2018 |
| INICC | International Nosocomial Infection Control Consortium (INICC) | Multiple countries, Asia | 2017–2020 |
| Jordan | Jordan University of Science and Technology, King Abdullah University Hospital | Jordan | 2019 |
| LAOS | Lao-Oxford-Mahosot Hospital-Wellcome Trust Research Unit | Laos | 2003–2019 |
| NeoOBS | Global Antibiotic Research and Development Partnership (GARDP), St. George's, University of London. NeoOBS Neonatal Sepsis Observational Study | Multiple countries, Asia | 2018–2020 |
| Pakistan | – | Pakistan | 2007–2019 |
| Pfizer | Pfizer ATLAS Programme Antimicrobial Resistance Surveillance | Multiple countries, Asia | 2004–2020 |
| Resistance_lit | Resistance Literature Review | Multiple countries, Asia | 2008–2016 |
| Sentry | SENTRY Antimicrobial Surveillance Program Data | Multiple countries, Asia | 2013–2018 |
| SGUL | St George's Hospital, University of London. GARPEC Project Bloodstream Infection Data | Multiple countries, Asia | 2016 |
| Smart_merck | Study for Monitoring Antibiotic Resistance and Trends | Multiple countries, Asia | 2002–2021 |
| Systematic_review_2024 | Systematic review | Multiple countries, Asia | 2000–2021 |
| Taiwan | National Institute of Infectious Diseases and Vaccinology (Taiwan) | Taiwan | 2007–2016 |
| Thailand | – | Thailand | 2017–2019 |
| Vietnam | Oxford University Clinical Research Unit. Ho Chi Minh City Hospital for Tropical Diseases Prospective Observational Study of Hospital-acquired Infections in ICU Patients | Vietnam | 2012–2016 |

### B Model Derivation

A two-compartment ODE model can be used to represent the general dynamics of colonisation with drug-resistant strains. This framework assumes that colonisation with drug-sensitive strains remains approximately constant over time, and so drug-sensitive colonisation is not explicitly modelled. The differential equations describing the dynamics of this two-compartment model are shown in Equation 1 and Equation 2. Here, the *X* represents uncolonised individuals susceptible to infection and *I* represents individuals currently infectious.

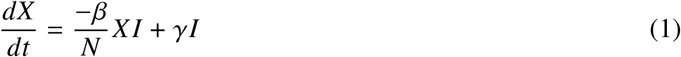

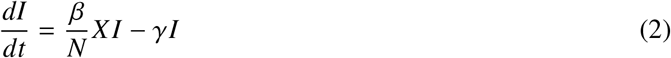

We assume a constancy of the population *N* to obtain a proportional value of the total population colonised with drug-resistant strains and can rewrite Equation 2 in terms of *I, β, γ* under this assumption.

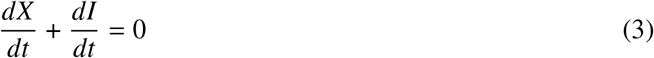

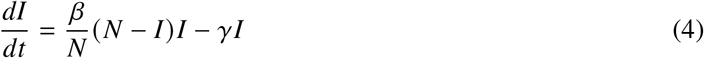

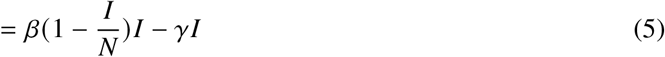

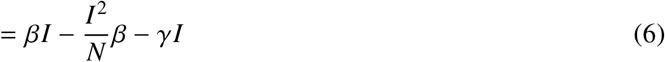

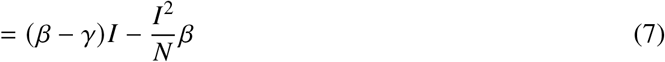

An analytical solution to the two-compartment model is the Verhulst equation, also known as the logistic growth function (see Equation 10). We use the logistic growth function as an analytical solution to this two-compartment model [44]. This equation can be expressed in terms of the initial proportion of resistance *P*_0_, a maximum limiting value for the population *K*, and the growth rate *r* (see in Equation 9) [45].

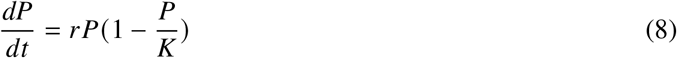

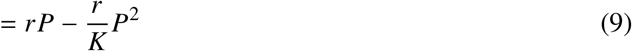

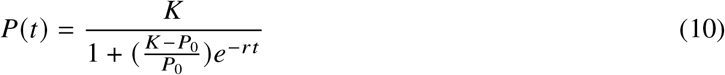

We restructure Equation 7 in terms of Equation 9 to find the growth rate *r* and carrying capacity *K* in terms of *β* and *γ*.

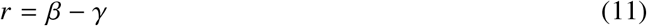

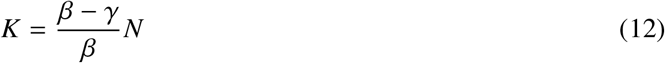

This allows us to express Equation 10 in terms of *β* and *γ*, as shown in Equation 13.

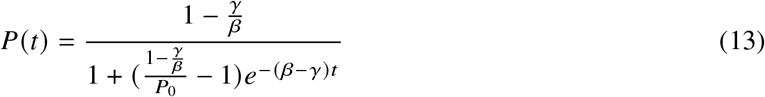

While this is undefined when *β* = *γ*, by applying L’Hôpital’s rule we find that 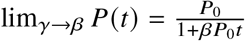.

### C Covariate Models

For models with covariates, the priors for *P*_0_ and *γ* remain the same as in the country-level effects model and the priors for *β* are specified below.

In Equation 14, *θ*_*k*_ is the coefficient of the covariate *k* and is set to have a weakly informative normal prior distribution. The covariate values are then multiplied by their estimated coefficient value, summed over the total number of covariates, and then added to their estimated country-level *β*_*c*_ term. The final value is then exponentially transformed.

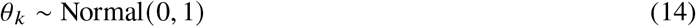

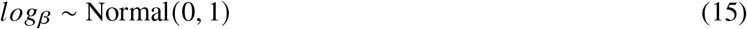

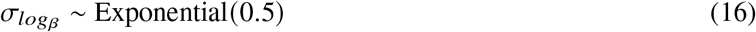

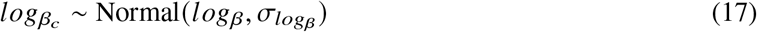

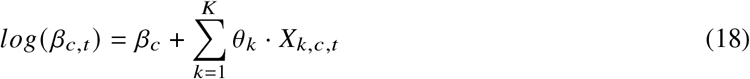

### D Spatial Effects

To incorporate spatial effects into the model, an initial approach used the intrinsic conditional auto-regressive (ICAR) component of the Besag-York-Mollié (BYM) model to smooth estimates across neighbouring countries and account for spatial interactions between pairs of countries [46–48]. We incorporate these spatial models following the implementation of BYM model in Stan [49, 50]. We add in parameter *ϕ* to encode the neighbour relationships in terms of the pairwise difference formula as shown in Equation 19. The pairwise difference formula is calculated between *i* and *j* if they are neighbours and under the condition that *i* cannot equal *j* . A soft sum-to-zero constraint is also applied to *ϕ* to soft-centers the mean close to zero and is shown in Equation 20 [49]. A precision term, *τ*_*ϕ*_, is added to estimate the standard deviation of *ϕ*, as shown in Equation 22. The spatial component, *σ*_*ϕ*_ * *ϕ*_*c,i*_, is added related to *I*_0_ and *β* as we expect neighbouring regions to have similar initial resistant rates and influence on transmission.

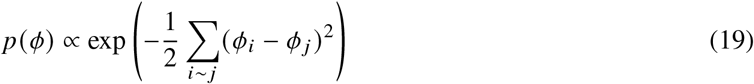

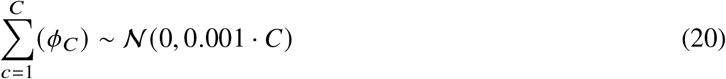

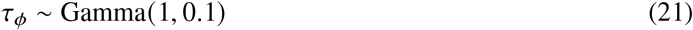

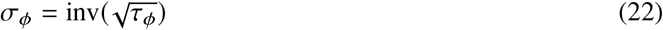

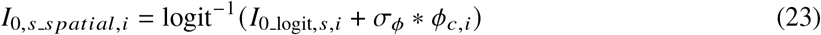

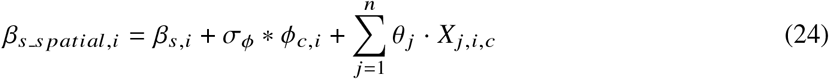

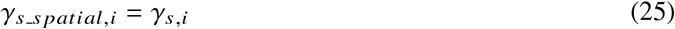

After testing spatial effects in terms of the BYM model, an alternate method was explored in terms of Gaussian process regression with the aim of reducing model run times and improving model convergence [35, 36].

To model the spatial relationship between countries, a kernel function can be used to define the covariance between countries based on distance. The radial basis kernel functions is shown as Equation 26 where *D* is the distance between countries, *ζ* ^2^ is the maximum covariance between countries, and *ρ*^2^ is the rate that covariance declines. This relationship is then used to estimate the a spatial intercept term for each country following methods from previous examples [35, 36].

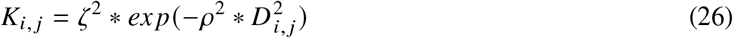

We incorporate the spatial effects intercept term,*k*, for *P*_0_, *β*, and *γ*. For *P*_0_, the spatial term is added to the population-level term and transformed to the inverse logit space as shown in Equation 27. For both *β* and *γ*, we add the spatial term in the log space and then exponentiated the sum of the spatial term and the population level parameter. The *β* and *γ* terms used in the final probability equation are shown in Equations 28 and 29.

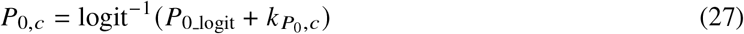

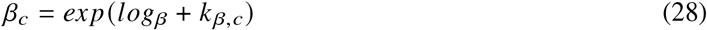

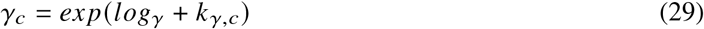

### E Country-level Priors

The prior distributions are shown in equations 30-39. Here, we have two options we explored for the initial resistance *P*_0_ prior. Option 1: We select a weakly informative normal prior distribution for *P*_0 logit_ since we do not have specific information on this parameter on a logit scale and having a hierarchical structure. Option 2: We select a beta(2,2) prior distribution for the initial resistance value for each country with no hierarchical structure. The first option was preferred for fitting models starting at the initial time point for each country. The second option was preferred for fitting models all from the same year and starting at previous years where data is sparse since it is computationally more efficient. For this analysis, we use the second option for the initial resistance prior as we are generating predictions from 2000 onwards and data is sparse. For *β* and *γ* we select an exponential prior distribution with a value of three since the value for these parameters have a higher probability density closer to 0 and should be positive. 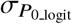, *σ*_*β*_, and *σ*_*γ*_ all have an exponential prior distribution with a value of 0.5 as we expect this value to be positive and expect to have a slightly higher probability density closer to 0 since we do not expect major differences between countries

for these parameters.

Prior distributions for the hierarchical model:

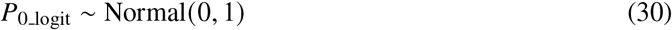

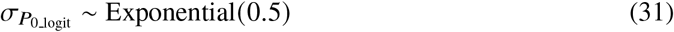

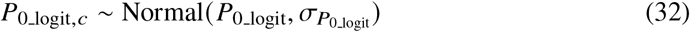

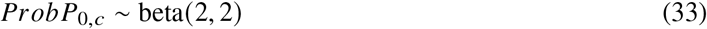

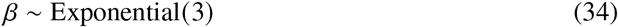

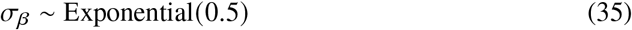

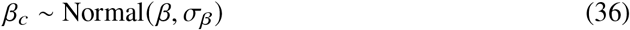

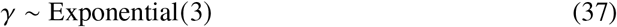

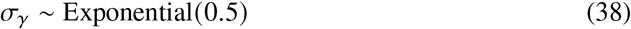

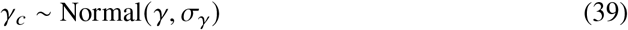

After defining these priors, we can define the *P*_0,*c*_, *β*_*c*_, and *γ*_*c*_ parameters.

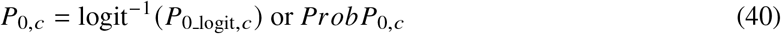

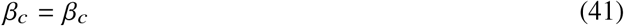

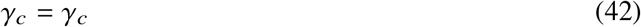

### F Data source-level Scaling

The prior distributions for the source-level parameters is shown. The models incorporating source-level scaling extend the country-level model and use the same priors as shown in the previous section for *β*_*c*_, *γ*_*c*_, and *P*_0,*c*_. Here, the source-dependent multiplicative factor η_*s*_ is set to be bounded between 0 and 1 to ensure positive constraints and with a beta distribution density concentrated near 1. The structure of this model shows that the underlying curve for each country should be similar and the variation between sources should be relatively small when scaling the underlying curve.

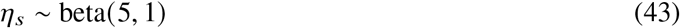

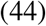

### G Resistance Trajectories

Resistance trajectories for each country across all pathogen-antibiotic combinations for Asia from 2000 to 2022.

**Fig. 4.**
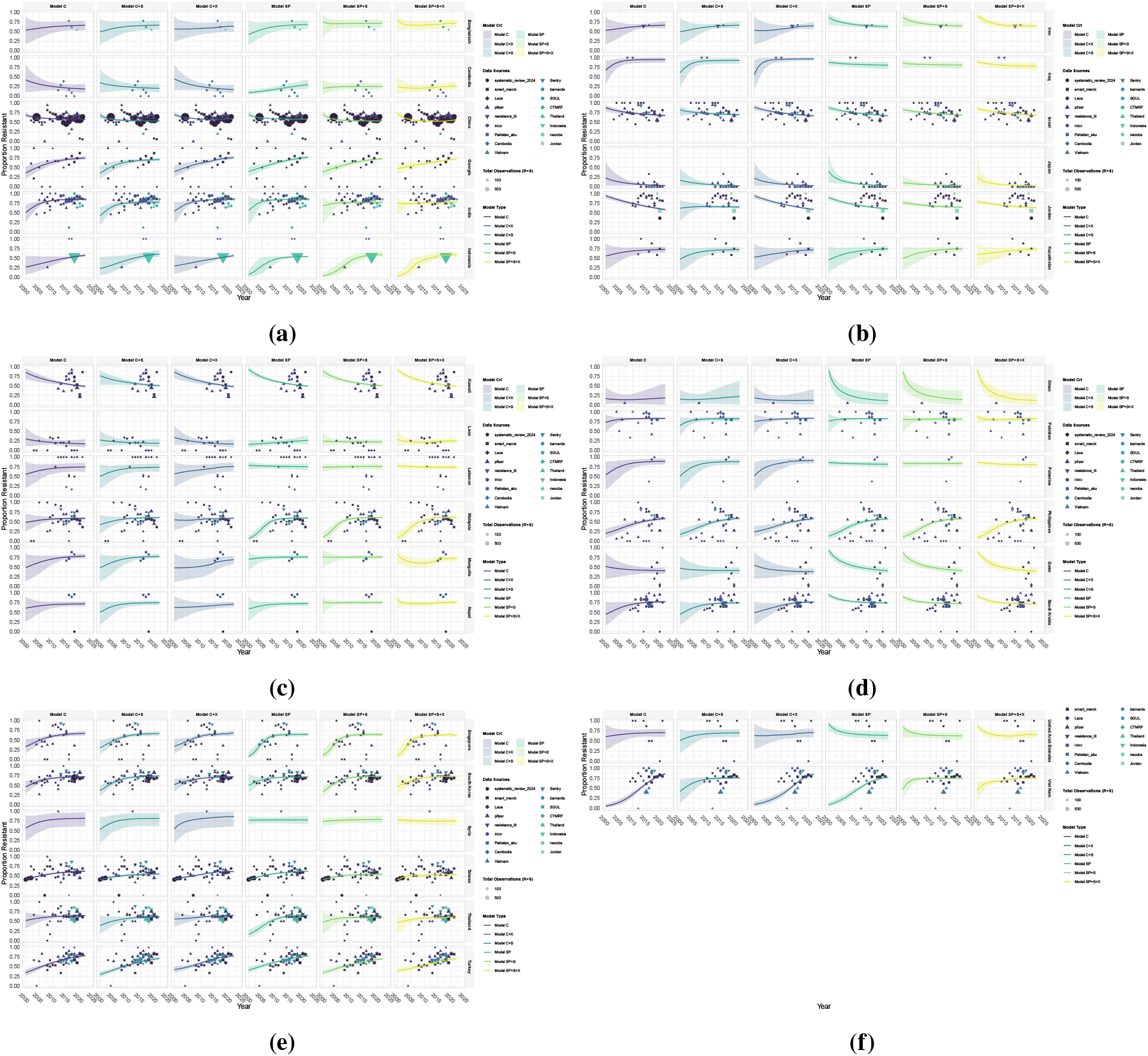
Individual model fits using *β γ, P*_0_ parameterisation of the logistic growth function for resistance of *Acinetobacter baumannii* to aminoglycosides model training for Asia. a) to e) shows the posterior predictions for the proportion of clinical isolates that are resistant to aminoglycosides for all countries and six model variants. Model C: Model with country-level random effects, C+X: Country-level random effects and covariates, C+S: Country-level random effects and source-level scaling, SP: Spatial model with Gaussian process regression, SP+S: Spatial model with Gaussian process regression and source-level scaling, SP+S+X: Spatial model with Gaussian process regression, source-level scaling, and covariates. Solid lines represent the mean posterior prediction and the shaded regions show the 95% credible intervals with the different colours corresponding to each model type. The points show the observed resistance data used for training the models.

**Fig. 5.**
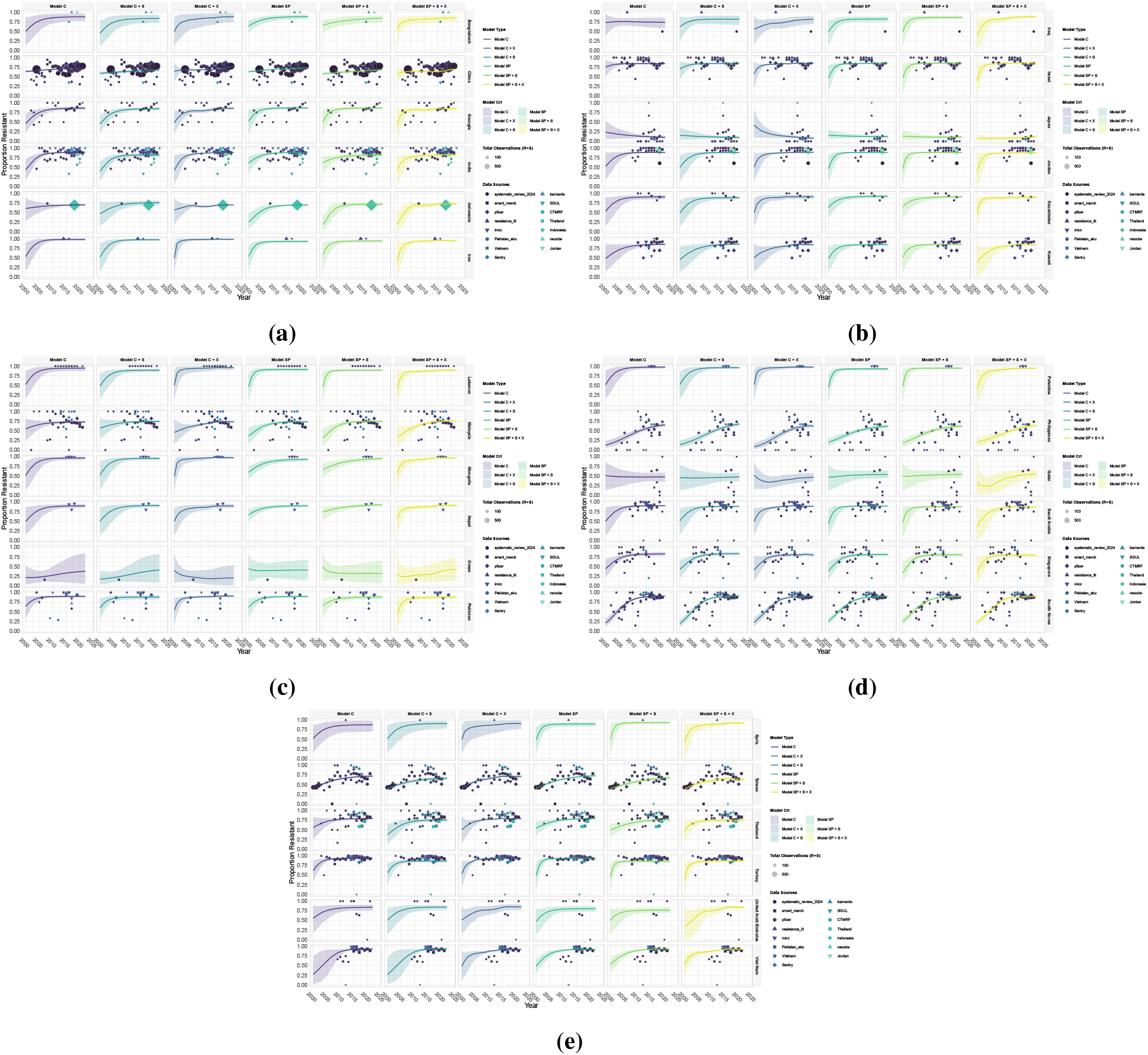
Individual model fits using *β γ, P*_0_ parameterisation of the logistic growth function for resistance of *Acinetobacter baumannii* to antipseudomonal penicillins model training for Asia. a) to e) shows the posterior predictions for the proportion of clinical isolates that are resistant to antipseudomonal penicillins for all countries and six model variants. See Fig 4 for model details. Solid lines represent the mean posterior prediction and the shaded regions show the 95% credible intervals with the different colours corresponding to each model type. The points show the observed resistance data used for training the models.

**Fig. 6.**
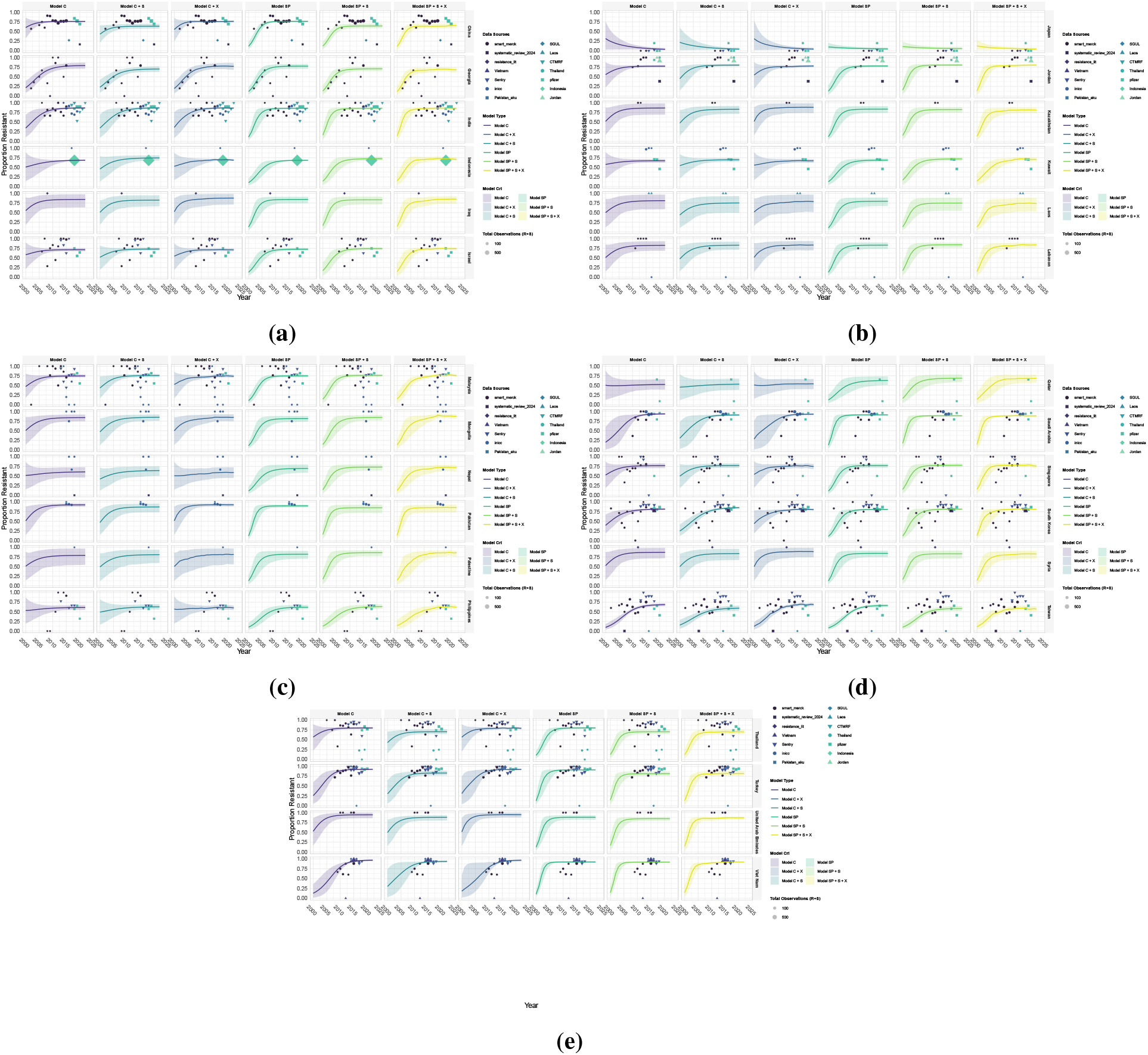
Individual model fits using *β γ, P*_0_ parameterisation of the logistic growth function for resistance of *Acinetobacter baumannii* to beta-lactams/beta-lactamase inhibitor combinations model training for Asia. a) to e) shows the posterior predictions for the proportion of clinical isolates that are resistant to beta-lactams/beta-lactamase inhibitor combinations for all countries and six model variants. See Fig 4 for model details. Solid lines represent the mean posterior prediction and the shaded regions show the 95% credible intervals with the different colours corresponding to each model type. The points show the observed resistance data used for training the models.

**Fig. 7.**
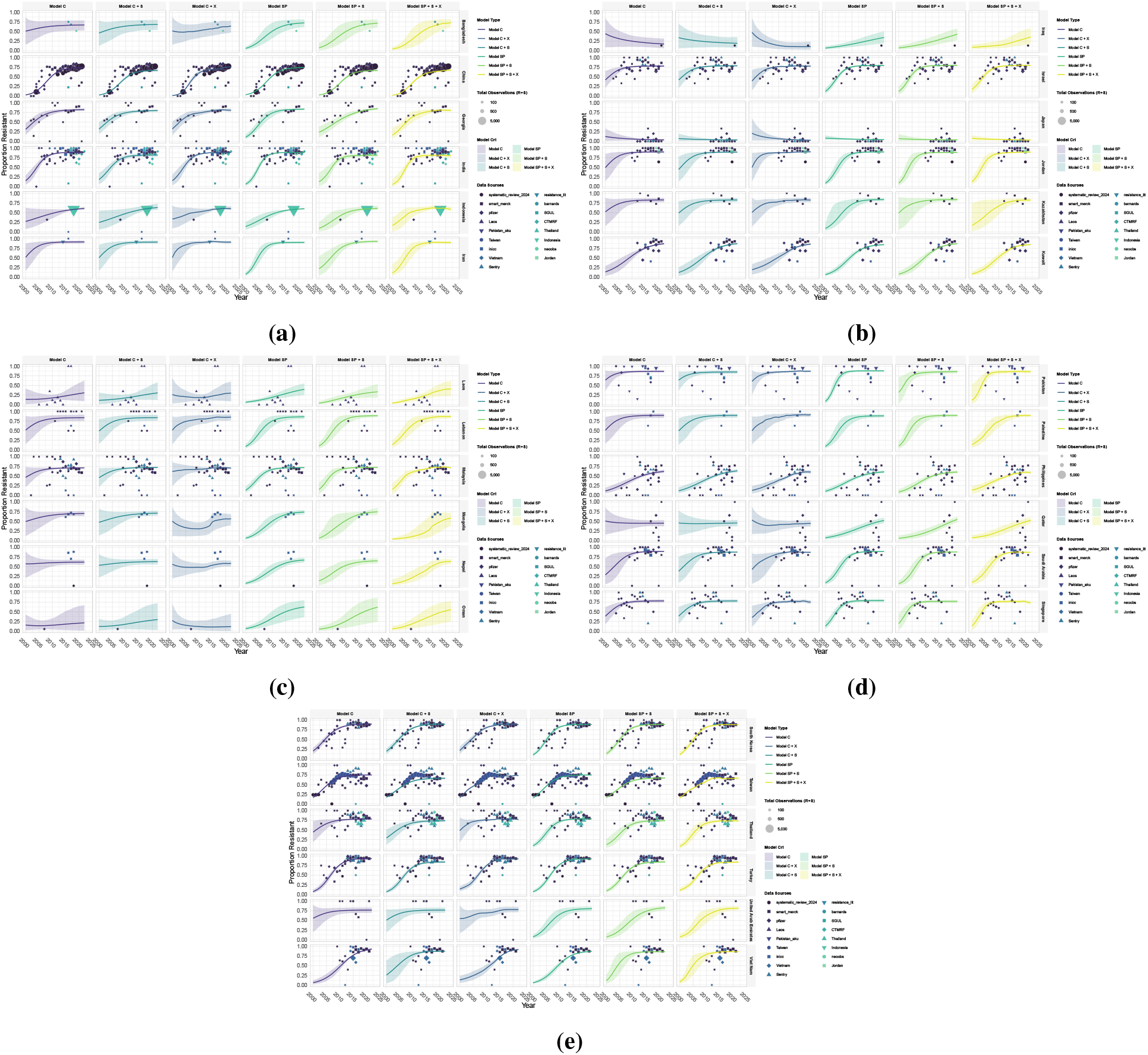
Individual model fits using *β γ, P*_0_ parameterisation of the logistic growth function for resistance of *Acinetobacter baumannii* to carbapenems model training for Asia. a) to e) shows the posterior predictions for the proportion of clinical isolates that are resistant to carbapenems for all countries and six model variants. See Fig 4 for model details. Solid lines represent the mean posterior prediction and the shaded regions show the 95% credible intervals with the different colours corresponding to each model type. The points show the observed resistance data used for training the models.

**Fig. 8.**
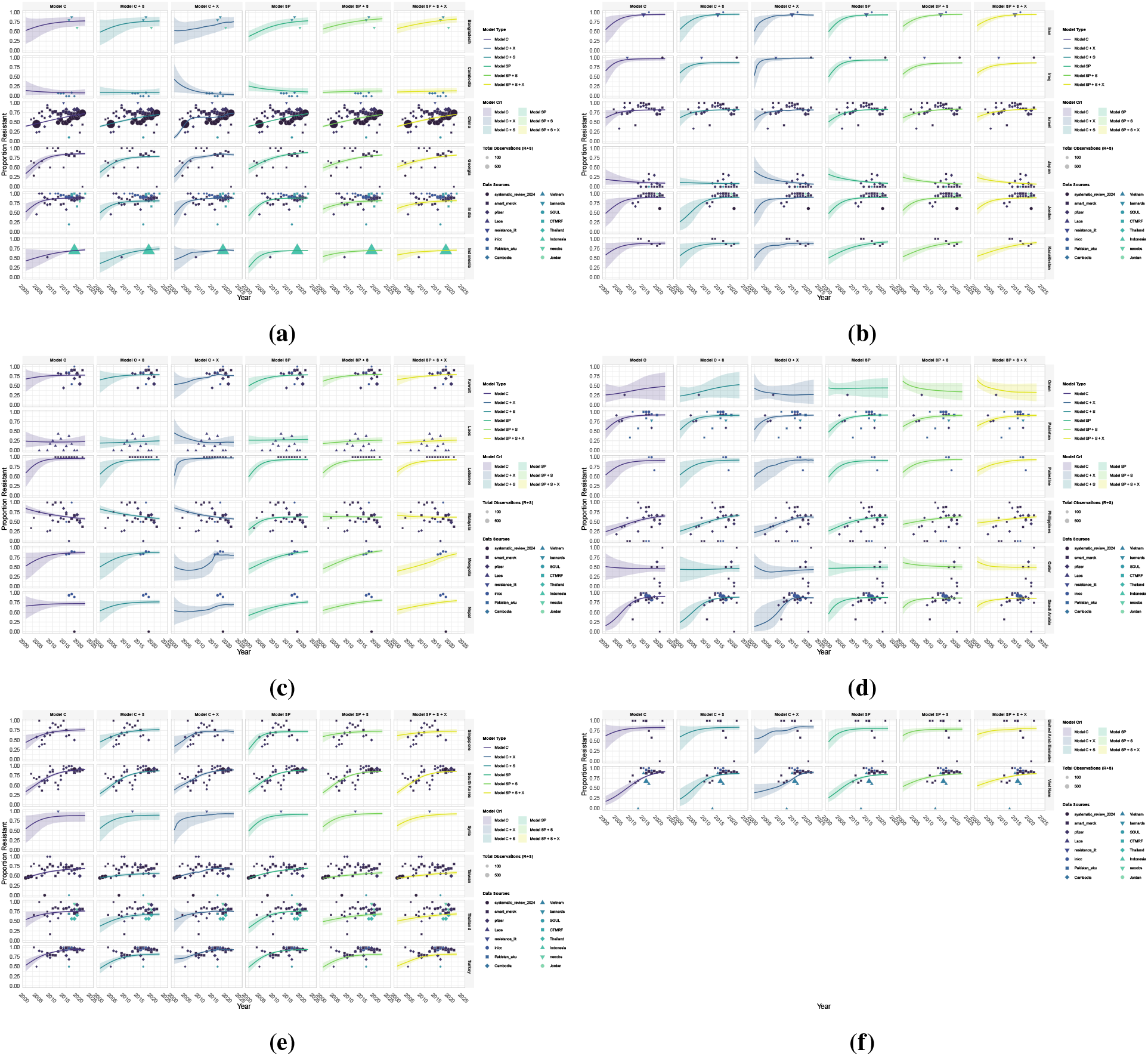
Individual model fits using *β γ, P*_0_ parameterisation of the logistic growth function for resistance of *Acinetobacter baumannii* to fluoroquinolones model training for Asia. a) to e) shows the posterior predictions for the proportion of clinical isolates that are resistant to fluoroquinolones for all countries and six model variants. See Fig 4 for model details. Solid lines represent the mean posterior prediction and the shaded regions show the 95% credible intervals with the different colours corresponding to each model type. The points show the observed resistance data used for training the models.

**Fig. 9.**
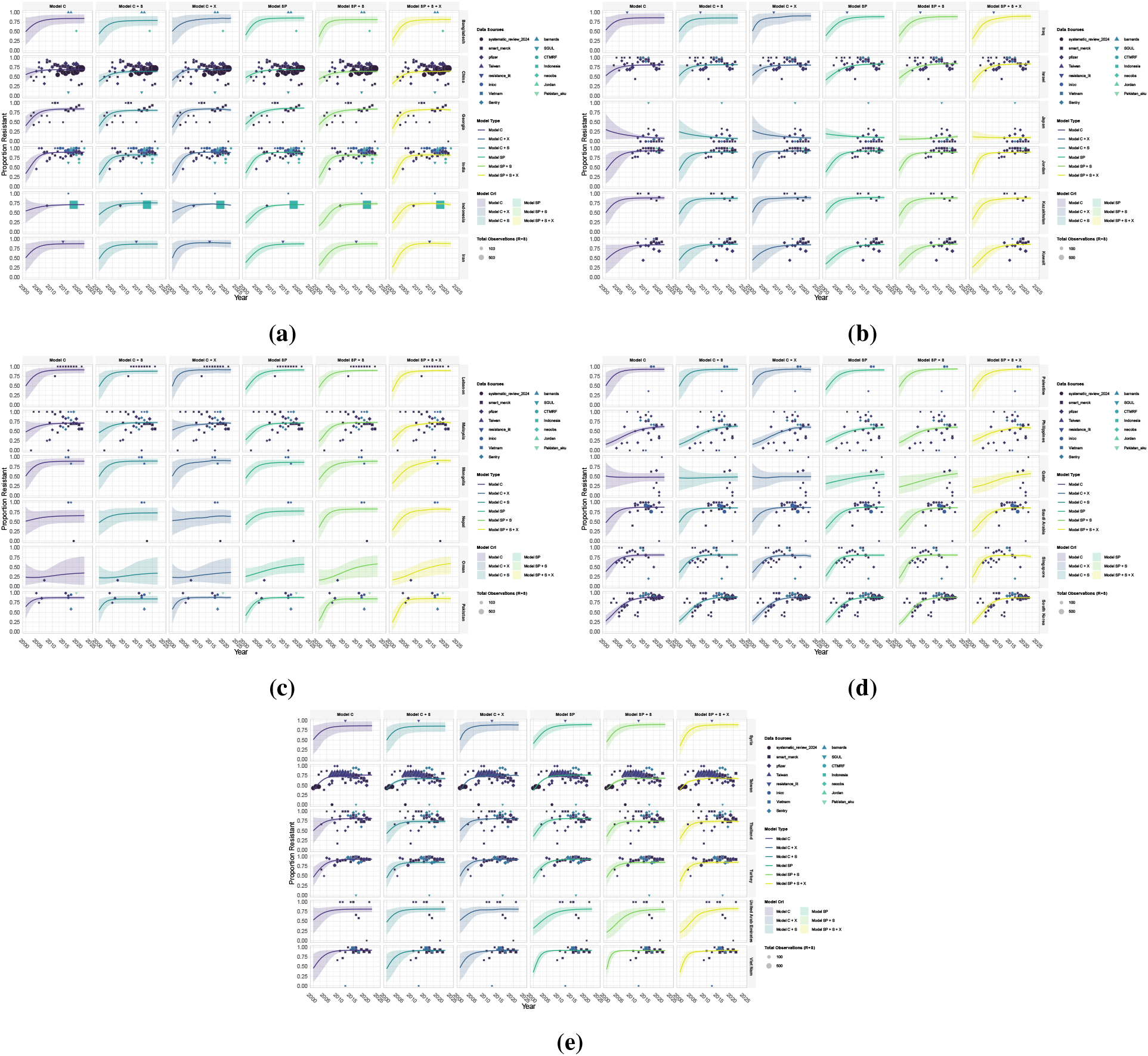
Individual model fits using *β γ, P*_0_ parameterisation of the logistic growth function for resistance of *Acinetobacter baumannii* to fourth-generation cephalosporins model training for Asia. a) to e) shows the posterior predictions for the proportion of clinical isolates that are resistant to fourth-generation cephalosporins for all countries and six model variants. See Fig 4 for model details. Solid lines represent the mean posterior prediction and the shaded regions show the 95% credible intervals with the different colours corresponding to each model type. The points show the observed resistance data used for training the models.

**Fig. 10.**
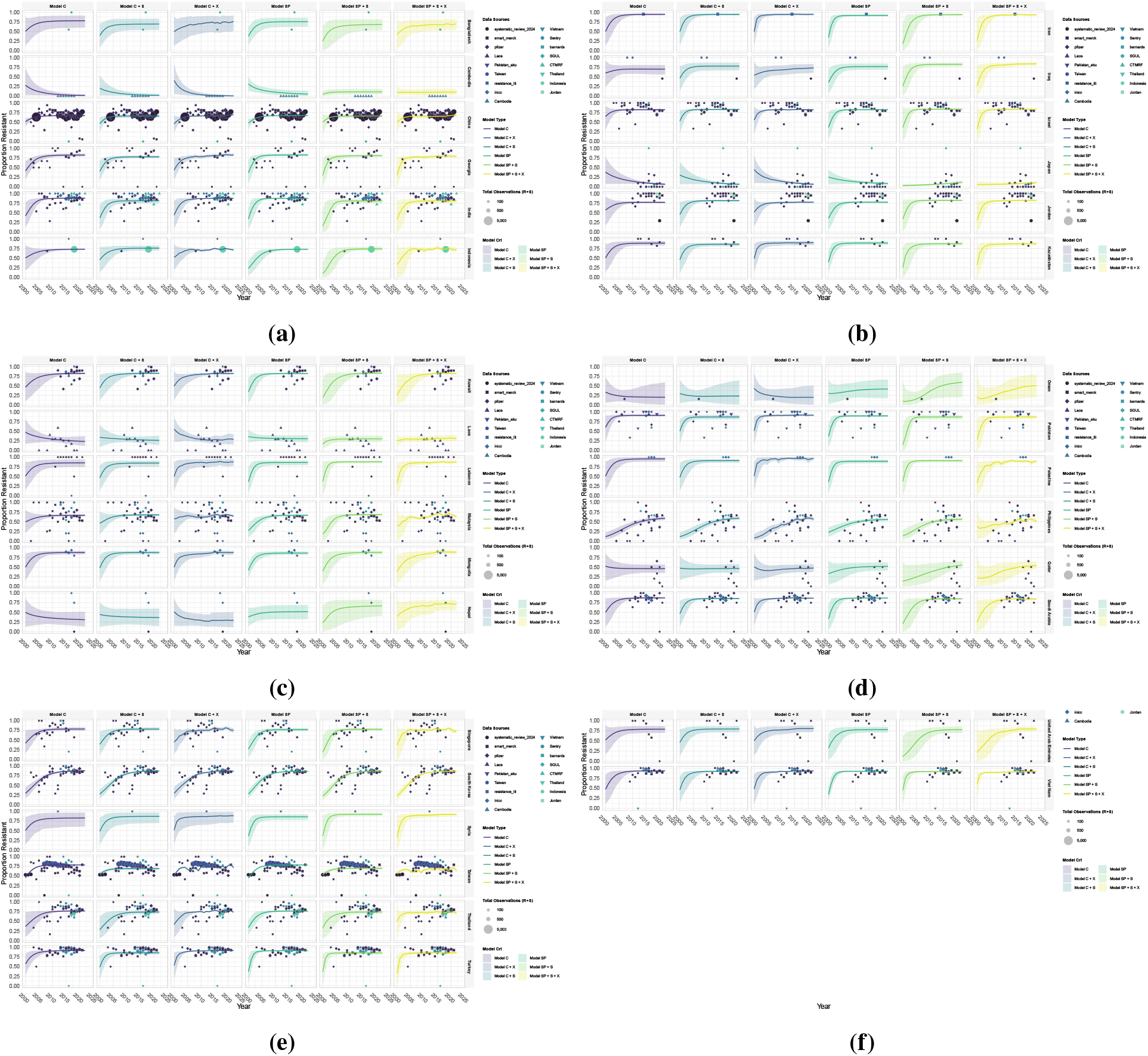
Individual model fits using *β γ, P*_0_ parameterisation of the logistic growth function for resistance of *Acinetobacter baumannii* to third-generation cephalosporins model training for Asia. a) to e) shows the posterior predictions for the proportion of clinical isolates that are resistant to third-generation cephalosporins for all countries and six model variants. See Fig 4 for model details. Solid lines represent the mean posterior prediction and the shaded regions show the 95% credible intervals with the different colours corresponding to each model type. The points show the observed resistance data used for training the models.

### H Long-term Saturation Levels

Long-term saturation levels estimated for each country for all pathogen-antibiotic combinations for Asia from 2000 to 2022.

**Fig. 11.**
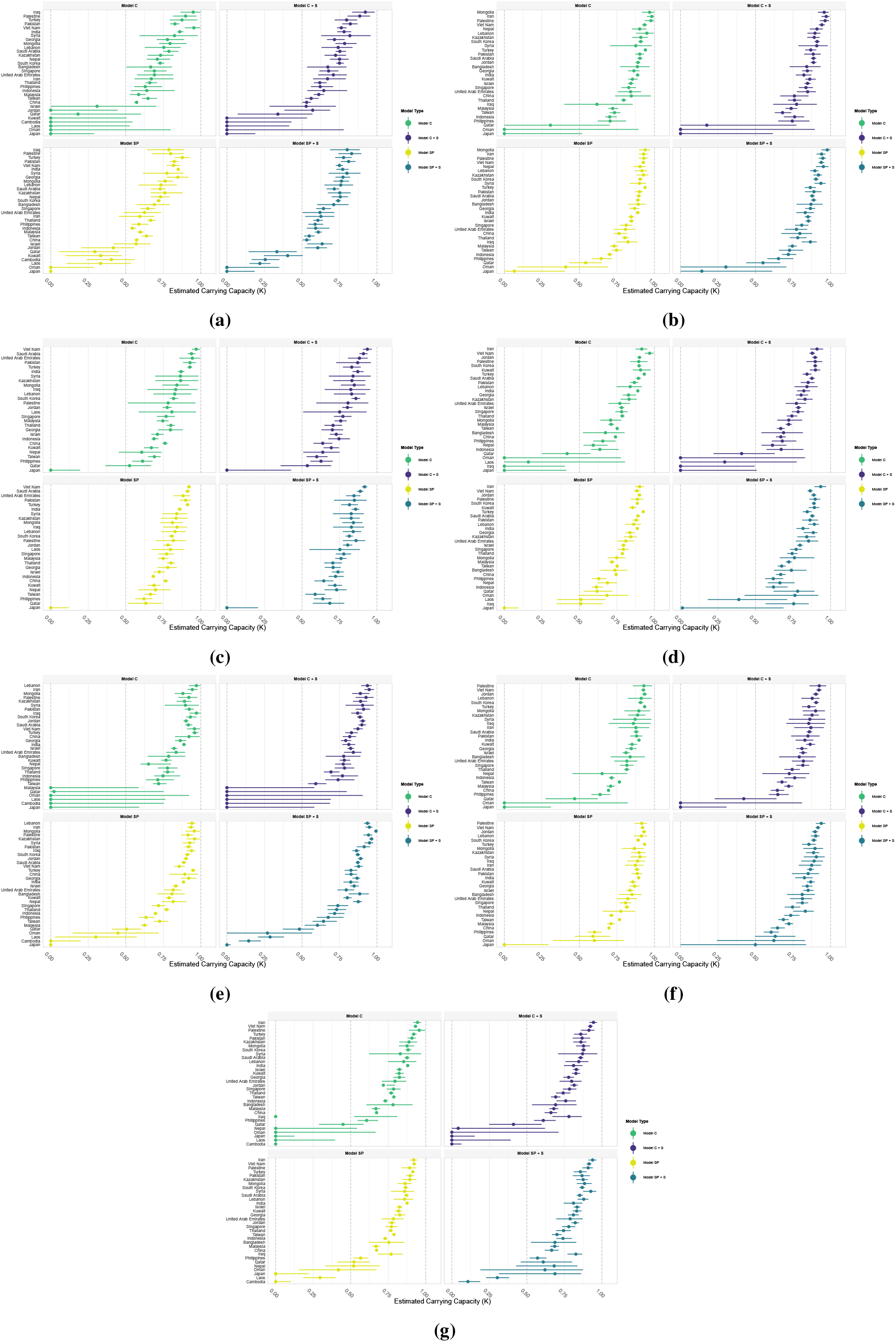
Long-term saturation resistance levels of *Acinetobacter baumannii* to antibiotic combinations for models trained for Asia. a) to g) show_1_s_8_ stabilisation values for aminoglycosides, antipseudomonal penicillins, beta-lactams/beta-lactamase inhibitor combinations, carbapenems, fluoroquinolones, fourth-generation cephalosporins, and third-generation cephalosporins, respectively. Points show posterior mean estimates with 95% credible intervals. Model C: Model with country-level random effects, C+S: Country-level random effects and source-level scaling, SP: Spatial model with Gaussian process regression, SP+S: Spatial model with Gaussian process regression and source-level scaling.

### I Covariate Model Predictions

The estimated covariate coefficients values are shown for models that incorporate covariates. These models include country-level random effects and covariates (C+X), and the spatial model with Gaussian process regression, source-level scaling, and covariates (SP+S+X). The covariates included are mean temperature, antibiotic consumption in terms of defined daily dose per 1000 inhabitant per day (DDD/1000), and out-of-pocket healthcare expenditures reported per year. Alternative covariates can be included in the model structure for extensions of this framework.

**Table 3.** Average coefficient value of covariates based on antibiotic class for mean temperature, antibiotic consumption in terms of defined daily dose per 1000 inhabitant per day (DDD/1000), and the out-of-pocket healthcare expenditures reported per year. The models incorporating covariates are country-level random effects and covariates (C+X), and the spatial model with Gaussian process regression, source-level scaling, and covariates (SP+S+X).

| Antibiotic Class/Subclass | Average Covariate Coefficient Estimated [95% CrI] |  |  |  |  |  |
| --- | --- | --- | --- | --- | --- | --- |
|  | Model C+X |  |  | Model SP+S+X |  |  |
|  | Temperature | DDD/1000 | OOP_hexp | Temperature | DDD/1000 | OOP_hexp |
| **Aminoglycosides** | 0.143 [-0.031, 0.309] | 0.210 [0.107, 0.322] | 0.009 [-0.117, 0.132] | -0.074 [-0.204, 0.127] | 0.251 [0.165, 0.337] | -0.028 [-0.133, 0.077] |
| **Antipseudomonal penicillins** | -0.082 [-0.224, 0.055] | 0.365 [0.284, 0.446] | 0.013 [-0.073, 0.095] | -0.010 [-0.159, 0.139] | 0.484 [0.391, 0.577] | 0.060 [-0.040, 0.159] |
| **Beta lactam/beta-lactamase inhibitor combinations** | -0.165 [-0.291, -0.040] | 0.057 [-0.088, 0.214] | 0.072 [-0.050, 0.190] | -0.147 [-0.267, -0.028] | 0.245 [0.131, 0.358] | 0.125 [0.012, 0.234] |
| **Carbapenems** | -0.199 [-0.300, -0.106] | 0.330 [0.249, 0.403] | 0.137 [-0.001, 0.273] | -0.192 [-0.287, -0.099] | 0.211 [0.112, 0.322] | 0.179 [0.062, 0.282] |
| **Fluoroquinolones** | -0.099 [-0.242, 0.042] | 0.511 [0.443, 0.581] | 0.186 [0.099, 0.274] | -0.044 [-0.195, 0.113] | 0.187 [0.094, 0.284] | 0.046 [-0.065, 0.156] |
| **Fourth-generation cephalosporins** | -0.128 [-0.210, -0.048] | 0.123 [0.020, 0.215] | 0.170 [0.067, 0.272] | -0.153 [-0.248, -0.059] | 0.170 [0.071, 0.268] | 0.189 [0.076, 0.298] |
| **Third-generation cephalosporins** | -0.412 [-0.504, -0.326] | 0.142 [0.073, 0.214] | 0.011 [-0.075, 0.107] | -0.353 [-0.442, -0.263] | 0.254 [0.166, 0.342] | 0.056 [-0.055, 0.177] |

### J Posteriors Densities from Simulated Data Model Fits

The models are fitted using simulated data with known model parameters to confirm parameter estimates are recoverable. The parameter posterior densities resulting from fitting simulated data with the country-level random effects model with a *β, γ, I*_0_ (initial proportion colonised with drug-resistant strains *P*_0_ in our model) parameterisation are shown. The resulting posterior densities are shown with the corresponding true parameter value used to generate data.

**Fig. 12.**
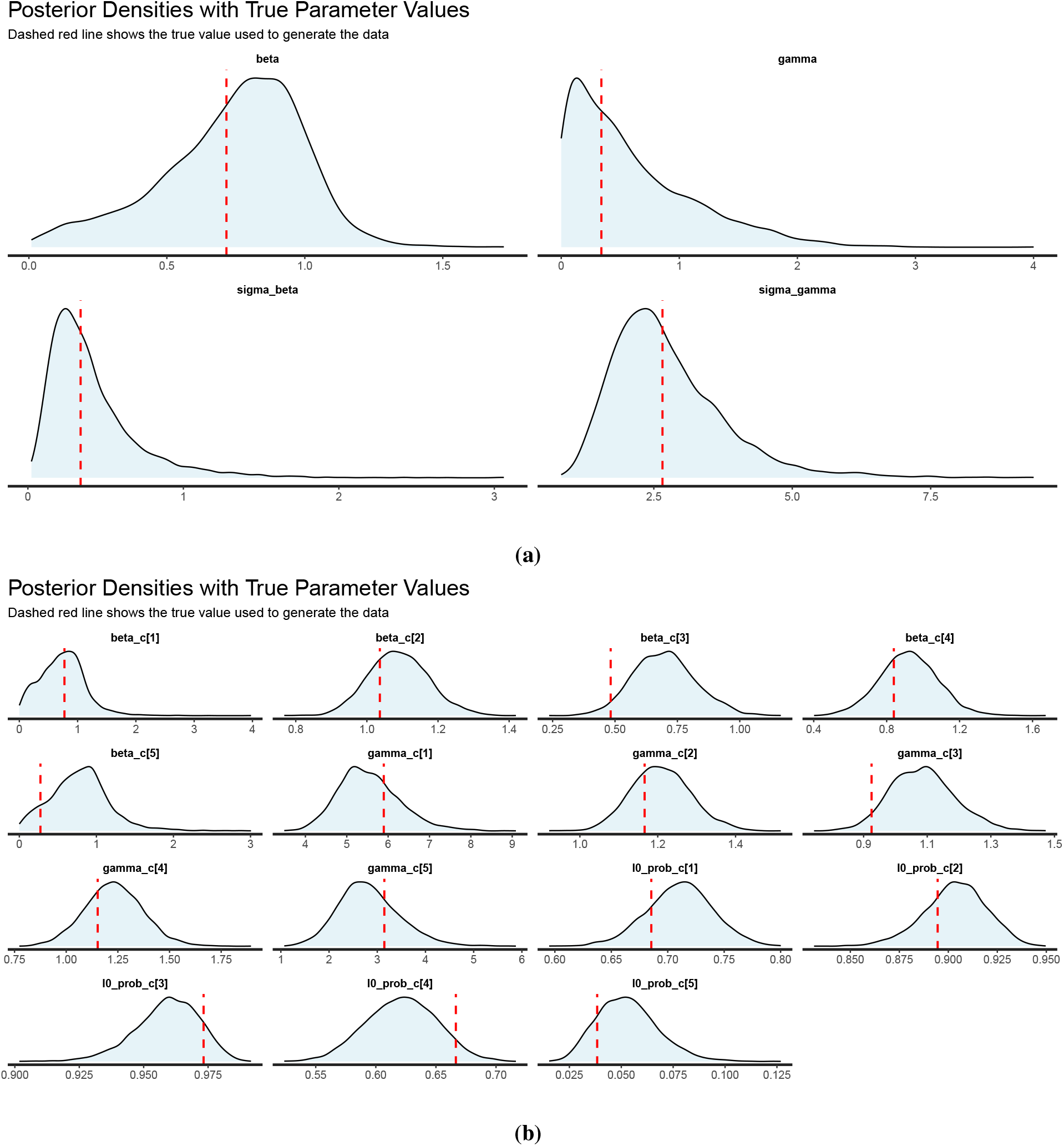
Posterior densities from data simulation using *β γ* parameterisation of the logistic growth function. a) shows the population level *β* and *γ* and b) shows the country-specific posterior densities recovered using the true parameter values shown by the dashed red line.

### K Model Trace and Pair Plots

Model convergence was checked through trace and pair plots generated after fitting models to *Acinetobacter baumannii* resistance data for each pathogen-antibiotic combination. Example pair and trace plots for carbapenems are shown.

**Fig. 13.**
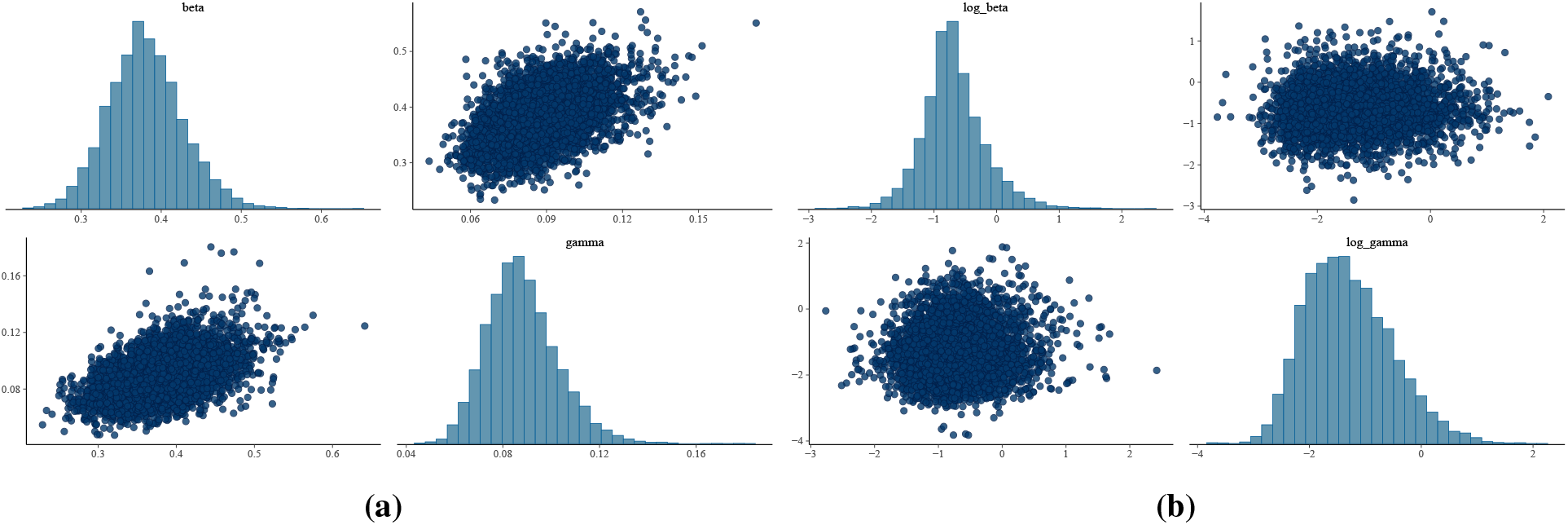
Pairs plot of *β* and *γ* the using *β γ, P*_0_ parameterisation of the logistic growth function for resistance of *Acinetobacter baumannii* and carbapenems model trained for Asia from 2000 to 2022. a) Shows pairs plots of country-level random effects b) pairs plot for base spatial-effects model

**Fig. 14.**
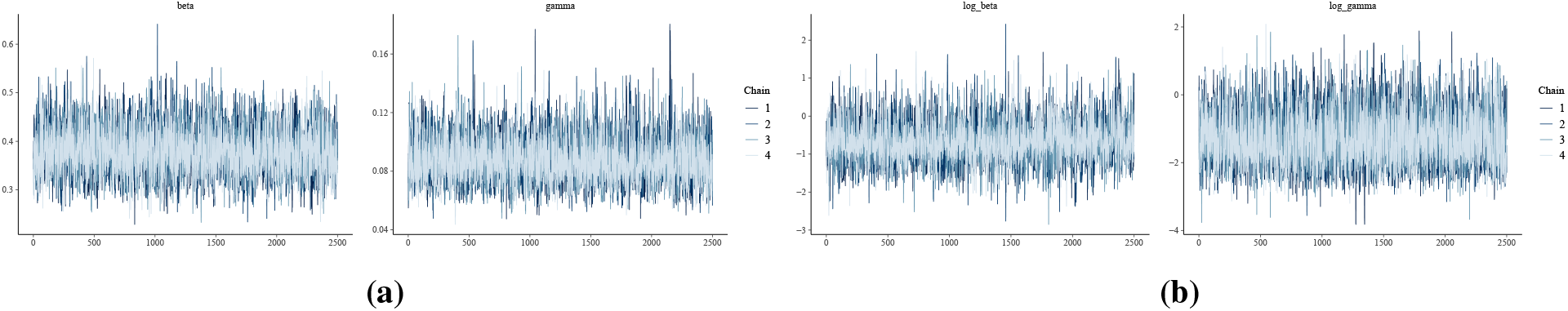
Trace plots of *β* and *γ* the using *β γ, P*_0_ parameterisation of the logistic growth function for resistance of *Acinetobacter baumannii* and carbapenems model trained for Asia from 2000 to 2022. a) Shows trace plots of the country-level random effects model b) trace plot for the spatial-effects model

